# Impact of mass oral cholera vaccination in an endemic area of the Democratic Republic of the Congo: a surveillance-based counterfactual modeling analysis

**DOI:** 10.64898/2026.09.17.26363298

**Authors:** Javier Perez-Saez, Espoir Bwenge Malembaka, Judith A Bouman, Patrick Musole Bugeme, Chloe Hutchins, Jules Jackson, Esperance Tshiwedi-Tsilabia, Juan Dent Hulse, Jaime Mufitini Saidi, Baron Bashige Rumedeka, Moïse Itongwa, Oliver Cumming, Esther German, Jean-Claude Kulondwa, Amy Dighe, Christy Clutter, Justin Lessler, Daniel T Leung, Karin Gallandat, Elizabeth C Lee, Placide Welo Okitayemba, Daniel Mukadi-Bamuleka, Jackie Knee, Andrew S Azman

**Author notes:** Correspondence: Andrew S Azman, 9 chemin des mines, 1202, Geneva, Switzerland.

## Abstract

**Background:** Oral cholera vaccines (OCVs) are a key component of cholera control recommended in cholera-endemic areas. Yet evidence of their population-level impact is limited, especially in Africa where most cholera deaths occur. Here, we estimate the impact of mass administration of OCVs in the cholera-endemic city of Uvira, Democratic Republic of the Congo.

**Methods:** We conducted enhanced cholera surveillance at the two official cholera treatment facilities in Uvira between Jan 2017 and Dec 2023, centered around a mass vaccination campaign that achieved 66% coverage with at least one dose of Euvichol Plus vaccine in 2020. We combined systematic rapid diagnostic case testing with repeated, representative population surveys capturing healthcare-seeking behavior, vaccination, population mobility, and antibody profiles to estimate seroincidence. We developed a Bayesian framework that integrates these data into an ensemble of mechanistic cholera transmission models that account for time-varying transmissibility, realistic immunity dynamics and loss of vaccination coverage to population turnover. OCV impact was assessed through ensemble counterfactual modeling of alternative vaccination scenarios.

**Findings:** We estimate that the 2020 mass vaccination averted 56% (95% Credible Interval: 34-81) of infections and deaths over the subsequent three years. This corresponds to 2,350 (median, 95% CrI: 890-8,960) averted facility-attended cases, and 44 (median, 95% CrI: 16-150) averted facility and community deaths. Vaccination of all of Uvira’s eligible population would have averted 64% (median, 95% CrI: 43-87) of cases and deaths, with a negative but limited influence of vaccine coverage loss because of population turnover (71% median, 95% CrI: 52-90 at half the population turnover).

**Interpretation:** Although mass vaccination averted a significant fraction of cholera cases that would have otherwise occurred in this endemic setting, imperfect coverage, population turnover, and high transmission rates contributed in offsetting the larger potential benefits of OCV. Successful cholera control in Uvira hinges on multisectorial approaches including provision of safe water and sanitation. Setting and communicating realistic expectations for mass vaccination programs in highly endemic areas is critical for maintaining confidence in the current generation of OCVs.

**Funding:** Gavi (M&E 9166 09 20 A16) and the Wellcome Trust (221688/Z/20/Z).

**Research in Context:** *Evidence before this study:* We searched PubMed for studies published between January 1, 2000 and June 2026 using the terms (“oral cholera vaccine” OR “OCV”) AND (“vaccine effectiveness” OR “vaccine impact” OR “indirect protection” OR “herd immunity” OR “population-level protection”). We also reviewed World Health Organisation (WHO) position papers and reports from the Global Task Force on Cholera Control (GTFCC). Inactivated oral cholera vaccines provide strong direct protection against symptomatic cholera, with randomized controlled trials and observational studies reporting effectiveness estimates typically exceeding 50% in the first three years after vaccination. In a previous study we showed that one-dose vaccine effectiveness was 52% 12-17 months after vaccination in Uvira, DRC. However, evidence on the population-level impact of preventive mass vaccination campaigns, which encompasses both direct and indirect protection, is thin. The only published estimates from clinical trials, both conducted in South Asia, reported overall protection of 37% over a 2-year period in Dhaka, Bangladesh and 75% over a 3-year period in Kolkata, India. No rigorous real-world estimates of mass OCV impact have been published from sub-Saharan Africa, where the vast majority of global cholera deaths occur. A major obstacle behind this gap is the lack of high-quality pre- and post-vaccination surveillance to support robust estimation of impact. The WHO and GTFCC have identified this evidence gap as a major research priority.

*Added value of this study:* To our knowledge, this is the first study to provide a rigorous, data-driven estimate of the population-level impact of a preventive mass OCV campaign in a cholera-endemic African setting. Drawing on 7 years of enhanced clinical surveillance, three population-representative household surveys measuring vaccine coverage, population mobility and health-care seeking behaviors, a serological survey, and a novel ensemble of compartmental transmission models, we estimated that the 2020 campaign in Uvira, DRC — which achieved approximately 66% coverage of at least one dose of Euvichol Plus vaccine — averted 56% (95% CrI: 34-81) of cholera cases and deaths over the subsequent three years. This corresponds to 2,350 (median, 95% CrI: 890-8,960) averted cases that would have sought care at official cholera treatment facilities, and 44 (median, 95% CrI: 16-150) averted deaths. Our transmission modeling framework also yielded new insights into cholera natural history in this endemic setting: symptomatic infection conferred strong and durable immunity (mean duration 6 years), while immunity following asymptomatic infection was substantially weaker. We further quantify how vaccination coverage and population turnover jointly determine vaccination program impact, providing a practical framework for informing future campaign design.

*Implications of all the available evidence:* Taken together with prior evidence from South Asia, our findings indicate that preventive mass OCV campaigns can avert a substantial fraction of cholera cases and deaths in highly endemic settings across different epidemiological contexts. Nonetheless, the resurgence of cholera a year after the vaccination campaign underscores that OCV campaigns alone are insufficient for sustained cholera control in similar settings due to high transmission rates and high population mobility. Investments in water and sanitation infrastructure remains essential.

## Introduction

Despite coordinated international efforts, the 7th cholera pandemic continues to pose a major global health threat as outbreaks and endemic transmission persist with little evidence of sustained improvement.^1^ While universal access to clean water and sanitation would largely eliminate cholera transmission, progress has been slow.^2^ Inactivated oral cholera vaccines (OCVs) serve as a critical tool to reduce cholera burden and are now an established component of global cholera prevention and control strategies.^3^

The World Health Organization (WHO) recommends using OCVs both reactively, for acute outbreaks and humanitarian crises, and preventively in endemic areas, though a global OCV shortage has forced most countries to prioritize reactive use.^4^ Several endemic countries, including the Democratic Republic of the Congo (DRC) and Bangladesh, have nonetheless secured doses for preventive campaigns in their highest-risk populations. Past modeling work had suggested that preventive mass vaccination at modest coverage may lead to cholera elimination in endemic settings.^5^ However, there is currently no empirical evidence for this as outbreaks tend to re-occur even in highly vaccinated endemic settings for reasons that remain unresolved. Therefore, the impact of a single mass campaign could be difficult to perceive directly: outbreaks continue, and their persistence may breed mistrust in the vaccine among communities and decision-makers even when many cases and deaths are being averted. Real-world evidence on the impact of mass vaccination could help set realistic expectations and guide resource allocation.

OCVs provide strong but waning direct protection against symptomatic disease, typically exceeding 50% in the first three years with estimates varying across settings.^6^ The population-level impact of a vaccination program reflects both this direct protection and the indirect protection conferred on unvaccinated and vaccinated people. ^7,8^ The only field estimates of this population-level effect come from two cluster randomized trials of two doses of OCV, both in South Asia: overall protection was 37% over a two year period in Dhaka, Bangladesh^9^ and 75% over three years in Kolkata, India.^10^ This large difference between estimates illustrates how population-level OCV impact may depend on multiple factors, including local epidemiology, vaccine coverage, the timing and spatial scale of mass vaccination as well as by population mobility.

Despite being identified as a major research priority by the Global Task Force on Cholera Control (GTFCC) and highlighted in the WHO position paper on oral cholera vaccines,^11,12^ to our knowledge there are no rigorous real-world estimates of population-level OCV impact beyond short-term outbreak contexts.^13^ This gap largely reflects the scarcity of endemic cholera settings with enhanced, long-term surveillance both before and after vaccination, as well as focused research studies to capture OCV coverage, healthcare seeking behavior, and asymptomatic infections.

Here, we use seven years of enhanced surveillance data in Uvira, a city in eastern DRC where cholera has been endemic since the late 1970s,^14^ combined with serial population-representative surveys, to estimate the overall impact of mass OCV campaigns using a suite of novel transmission models. We then use this framework to quantify the number of cases and deaths averted by OCV using counterfactual simulations. We further examine how vaccination program quality and population mobility shape the impact of mass vaccination and outline key lessons for future cholera vaccination programs.

## Methods

### Cholera surveillance in Uvira, DRC

Cholera surveillance is conducted through the two health facilities officially authorized to treat cholera patients free of charge in the city: the main cholera treatment center (CTC) located within the Uvira General Referral Hospital and a cholera treatment unit opened on July 16, 2019 at the Kalundu CEPAC health center (hereafter “the CTCs”). A suspected cholera case is defined as any individual aged at least 12 months presenting with three or more episodes of acute, non-bloody diarrhea (hereafter “acute watery diarrhea” [AWD]), within the 24 hours prior to CTC admission.

From August 2018, socio-demographic characteristics, clinical manifestations, and additional information recorded in the Ministry of Health register were collected using structured electronic questionnaires. Starting August 2021, vaccination status was systematically ascertained, through self-report, supported by structured questions and visual aids displaying OCV packaging, vaccination cards, and images of people taking oral vaccine doses.

Suspected cases were tested using the Crystal VC Rapid Dipstick test (O1/O139 or O1-only; Arkray Healthcare Pvt Ltd, Gujarat, India).^15^ Rectal swab specimens were collected by trained lab technicians or nurses, enriched in alkaline peptone water for six hours or longer depending on the time of patient admission, and then tested at the bedside for cases admitted before March 27, 2022, and in the onsite laboratory for cases admitted thereafter. Testing was not exhaustive during some periods due to logistical constraints, but reached close to 100%, except during periods of RDT stockouts (Figure 1).

**Figure 1.**
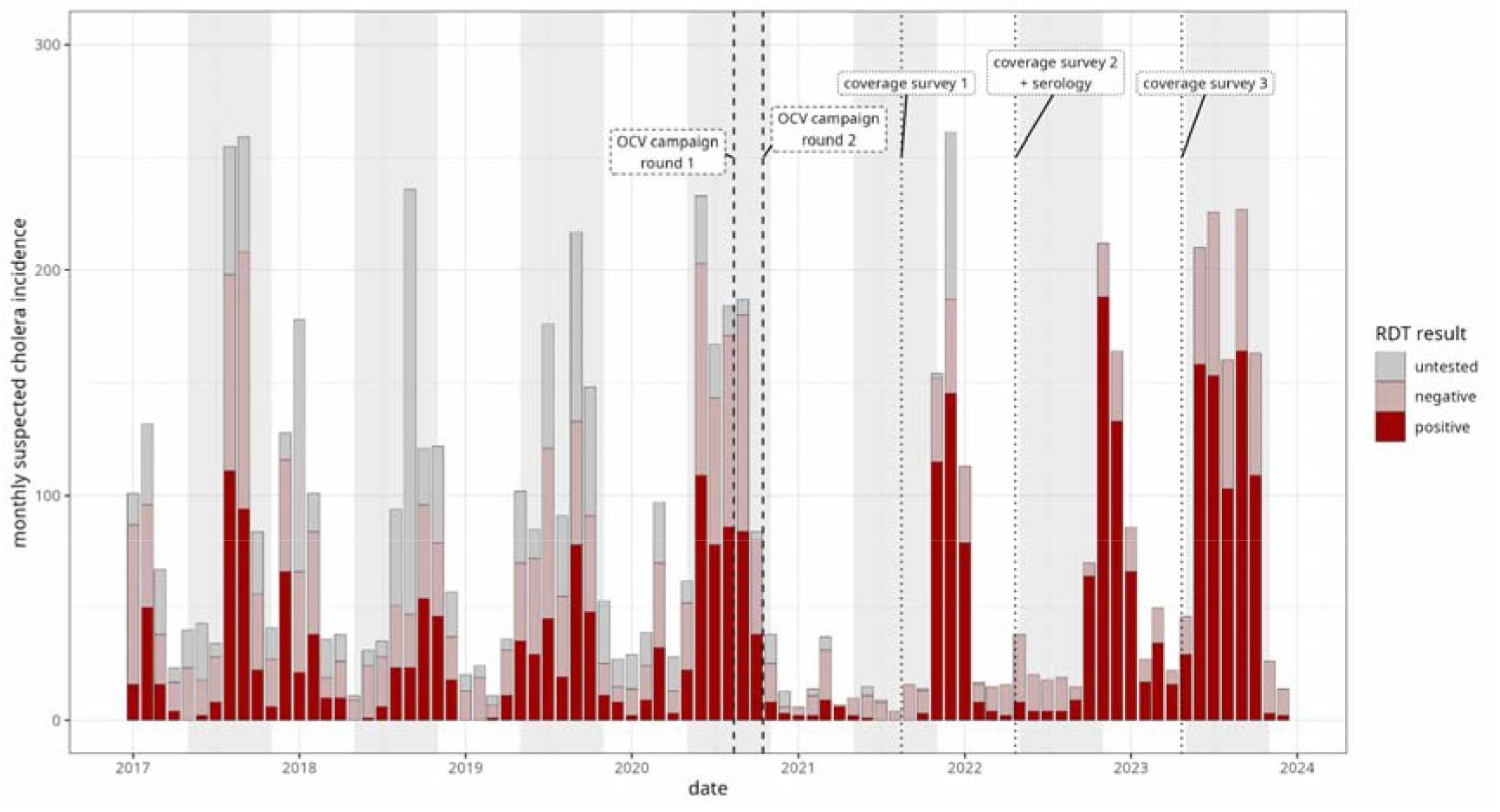
Monthly suspected cholera case counts in Uvira, DRC. Colors indicate RDT test results. Vertical lines indicate the midpoints of OCV campaign rounds (dashed lines) and of representative household surveys (dotted). Some suspected cases (in dark grey) were not tested due to stockouts of tests. Gray bands indicate months of typical high cholera incidence in the pre-vaccination period (May-October), referred to as the “cholera season” (Supplementary Figure S2).

### OCV campaigns

In response to severe flooding in Uvira in April 2020, the DRC Ministry of Health organized a mass oral cholera vaccination campaign using Euvichol-Plus (Eubiologics, Seoul, South Korea) ^16^. The campaign targeted all individuals aged ≥1 year living in the Uvira health zone, including pregnant women. The first round took place from July 29 to August 8, and the second from September 28 to October 5, 2020. Vaccination was delivered door-to-door for five days, followed by fixed-post vaccination at more than 22 sites, including health facilities throughout the community. Individuals were able to receive the vaccine in the second round regardless of whether they received one in the first round.

### Household surveys

We conducted three representative household-based surveys during the study period: the first in August 2021 (~11 months after the second vaccination campaign round), the second in April–May 2022 (~19 months after vaccination), and the third survey in April–May 2023 (~ 30 months after vaccination) as previously described.^16^ New samples were drawn for each survey round, using a random spatial sampling. In all surveys, vaccination status was assessed using the same approach as in clinical surveillance. The first survey included 2,288 individuals from 382 households. In the second survey, we enrolled 3583 individuals from 622 households, of which 2,376 individuals provided blood samples. In the third survey, we enrolled 2,864 people from 429 households. Through these surveys, we have previously estimated that 66% (95% CI 59-74) of the population received at least one dose of OCV in the campaign (41% with 2 doses).^16^ A vaccine effectiveness study conducted in Uvira over the same time period illustrated comparable protection between age groups from one dose over a three year period, with similar protection to previous 2-dose estimates.^16,17^ We also asked questions about household composition and household member migration, for which we used the 2022 survey to set priors on population-level migration rates (See Supplementary Material Section S3.2).

### Serology and seroincidence estimation

Serum was separated from whole blood on the date of sample collection and stored at −80C before being shipped to the University of Utah, where they were tested for IgG antibody responses to the O-Specific Polysaccharide (OSP, Inaba and Ogawa) and the B-subunit of the cholera toxin (CTB) following previously published protocols with a multiplex bead-based assay (Magpix, DiaSorin, Austin, Texas).^18^ More detailed methods are shown in Supplementary Material Section S1.

We used previously fit seroincidence models to classify each person’s serum antibody profile, or net median fluorescent index for each antibody target, as having been last infected in the past 200 days or not (200-day seroincidence model).^18^ We then adjusted these for the average sensitivity (47%) and specificity (95%) of the model using the Rogan-Gladen estimator^19^ to get an estimate of the cumulative incidence of infections (irrespective of symptoms) in the past 200 days. While vaccination elicits a strong immune response to these antigens, previous work has shown that the antibody kinetics of vaccinated individuals are indistinguishable from that of unvaccinated individuals a few months after vaccination, thus enabling robust seroincidence estimation.^20^

### Modeling analysis

The primary aim of this modeling analysis was to estimate the number of cases and deaths averted by the 2020 OCV campaign in Uvira. To this end, we fit a suite of five deterministic cholera transmission models with alternative representations of post-infection and vaccination immune dynamics. An overview of models is given in Supplementary Figure S1, and Supplementary Tables S2 (model features) and S3 (parameter definitions).

The simplest transmission models consisted of Susceptible Infected Recovered (SIR)-type models with time-varying transmission rates,^21^ thus allowing for cholera incidence seasonality (see wavelet analysis in Supplementary Material Section S2), an asymptomatic compartment, demographics, at least one-dose vaccination and leaky immunity^22^ (SIAR model). In the model, symptomatic infections represent the whole spectrum of severity, from mild to severe diarrhoea. We then expanded this model to account for differential immunity following symptomatic or asymptomatic infection^23^ (“two-path” model), and its expansion to allow for boosting of immunity through “unsuccessful” bacterial infections^22^ (“two-path-boost” model). For these two latter models, we considered alternatives where asymptomatic infections after vaccination lead to short-lived immunity, thus mirroring natural infections (“vaccine-mirrored”), or one where post-vaccination infections lead to long-lasting immunity regardless of symptoms.

Model outputs were fit to six distinct data streams: i) weekly incidence of suspected cholera cases, ii) weekly counts of RDT-positive tests among tested suspected cases, iii) weekly number of vaccinated cases among RDT-positive cases, iv) total population size in Uvira (annual Uvira City Council Official Population Estimates), v) vaccination coverage in the three representative household surveys, and vi) 200-day seroincidence from the 2022 serological survey. For i) we used a negative binomial model. We note that we modelled true cholera incidence, not just RDT-positive incidence, and in ii) we accounted for the imperfect performance of RDTs using meta-analysis estimates of sensitivity and specificity.^24^ We report main parameter estimates for the best-fitting model using pareto-smoothed importance sampling leave-one-out as implemented in the loo R package.^25^

We then used a model ensemble to assess OCV impact under counter-factual scenarios of initial vaccine coverage and population turnover, including one with no vaccination and the observed population turnover (“primary counterfactual scenario”). The model ensemble was built by stacking posterior counterfactual draws from the suite of model structures, weighted by stacking model weights.^26^ OCV impact was computed using four outcomes: the total number of cholera infections (asymptomatic and symptomatic) (“infections”), the number of symptomatic infections (“symptomatic infections”), the number of symptomatic infections that seek care in one of the cholera treatment centers (“facility-attended cases”), and the number of cholera deaths occurring either in the cholera treatment centers (“facility deaths”) or in the community (“community deaths”). Based on case fatality risk (CFR) estimates for medically-attended cholera in Uvira and counts of community deaths,^27^ we assumed a facility CFR to 0.8%, and a community CFR equal to 0.02%, which accounts for the fact that the vast majority of community cases in endemic areas are mild ^28^. We performed sensitivity analyses on key modeling assumptions, first on potential changes in health seeking behavior during the study period, and second on potential mis-estimation of seroincidence from our cross-sectional model (details in Supplementary Material Section 3.5).

Analyses were performed with R (version 4.5.2) and the Stan probabilistic programming language (cmdstan version 2.35). Data and code needed to reproduce primary analyses are available at https://github.com/GenevaIDD/uvira-ocv-impact.

## Results

Between January 1, 2017 and December 15, 2023, we recorded 6,939 suspected cholera cases in the cholera treatment facilities of Uvira, of whom 79.8% (5,540/6,939) were tested with RDT and 55.0% (3,050/5,540) were positive for *V. cholerae* O1 (Figure 1). RDT-positive cholera cases were observed year-round, although most occurred between the months of May and December, with peaks in May to October (Supplementary Figure S2). When considering cholera-season based years (1-April through 31-March each year), the average annual suspected case incidence was 4.2 per 1,000 per year in the pre-vaccination period, with 45.1% testing positive by RDT (1.9 confirmed cases/1,000/year). Cholera incidence declined dramatically in the cholera season after vaccination (2020-2021), a departure from the dominant yearly seasonal pattern of the pre-vaccination era (Supplementary Figure S2). During the first post-vaccination cholera season, suspected cholera incidence decreased by 76%, to 1.0 per 1,000 per year with RDT-positivity decreasing to 31.6%, and confirmed the cholera incidence rate decreasing by 84%, to 0.3 per 1,000. However, this lull ended in October 2021 with large annual outbreaks during the three following cholera seasons, resulting in an average post-vaccination (Nov 2020-Dec 2023) RDT-positive incidence similar to that of the pre-vaccination period (3.8 cases per 1,000).

In order to develop counter-factual post-vaccination epidemic trajectories, we fit a suite of models that differ in the way they represent immunity dynamics after infection and vaccination (see Methods section “Modeling analysis”). Through formal model comparison, models that allowed for differences in immunity by disease severity were favored (Supplementary Table S3). The best fitting model (Supplementary Table S4) was able to accurately reproduce key dimensions of the data, including suspected cholera incidence, RDT positivity, vaccine coverage over time and proportion of RDT-positive cases who were vaccinated (Figure 2). The model also captured changes in the population size of Uvira and the 200-day seroincidence estimate from our representative serosurvey (37.9% in March 2022) (Supplementary Figure S4).

**Figure 2.**
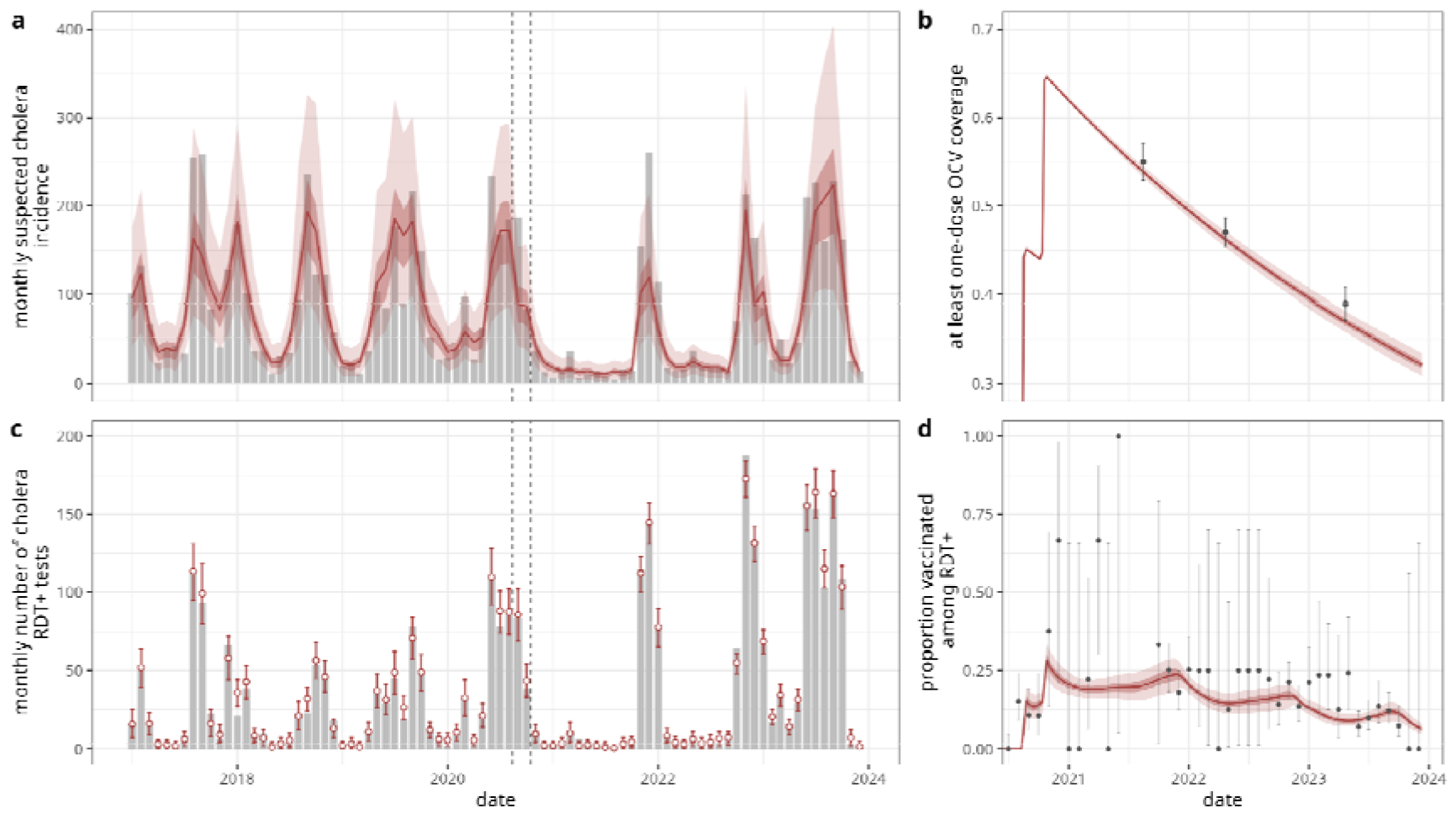
Model fidelity overview. Comparison of modeling outputs (red) against observations (gray) for a) monthly suspected cholera incidence, b) at least one-dose OCV coverage, c) monthly number of RDT-positive tests, and d) proportion of vaccinated individuals among RDT-positives. Vertical dashed lines indicate the midpoints of OCV campaign rounds 1 and 2. OCV coverage was obtained from representative household surveys as previously described.^16^ Modeled estimates correspond to the stacked model outputs, with lines and points indicating the mean, and bars and ribbons the 95% credible intervals of 2,000 posterior draws. Points and bars in black in panels b) and d) indicate the mean and the 95% binomial confidence intervals of the data.

We first report on inferred natural history parameters to contextualize our vaccine impact estimates. From our best fitting model, we infer that for every true symptomatic cholera infection, spanning the range of clinical severity, there are another 3.8 (95% CrI: 3.1-4.9) asymptomatic infections, and that symptomatic infections are 6.5 times (95% CrI: 2.5-29.6) more infectious than asymptomatic infections (Supplementary Figure S5). After a symptomatic infection or at-least one vaccine dose, we estimate an average duration of immunity of 6.1 years (95% CrI: 4.8-8.3) in the absence of boosting through re-exposures, which would be extremely challenging to measure directly in cholera-endemic settings.

When compared to a fully susceptible population, this equates to 84% (95% CrI: 80-87) protection against any type of infection after 1 year, and 60% (95% CrI: 53-69) protection after 3 years (Supplementary Figure S6). On the other hand, immunity from asymptomatic infection conferred 83% (95% CrI: 74-91) protection at the time of infection and lasted only 16.2 weeks on average (95% CrI: 7.0-33.4), with 16% (95% CrI: 2-37) protection after 6 months. In terms of population immunity levels, we find that most individuals in Uvira had some level of immunity from a previous infection at the start of the study period (93%, 95% CrI: 86-99). We also infer that incidence in Uvira is driven by high transmission rates with large seasonal fluctuations, with an average basic reproduction number (R_0_) of 9.0 (95% CrI: 6.1-12.1) ranging between a minimum of 2.8 (95% CrI: 1.4-4.7) and maximum of 33.0 (95% CrI: 18.5-54.0) across the study period (Supplementary Figure S6). All models showed large peak R_0_ values well above 10, except for the simplest SIAR formulation, which had markedly poorer fit to the data (Supplementary Figure S7, Supplementary Table S4).

We assessed the impact of OCV through counterfactual analysis using an ensemble of four models, excluding the simplest SIAR model because of unrealistic fit values in the 2022-2023 season outbreak (Supplementary Table S4, Supplementary Figure S9). We inferred that the 2020 OCV campaign averted 56% (95% CrI: 34-81) of infections and deaths in the three years after vaccination (Figure 3), with 79% (95% CrI: 67-94) of cases averted in the first year, 45% (95% CrI: 28-67) in the second and 48% (95% CrI: 11-77) in the third (Supplementary Table S6). The OCV campaign reduced the average annual symptomatic infection rate from 250 (95% CrI: 130-560) infections/1,000 person/year to 100 (95% CrI: 80-110) infections/1,000 person/year. This translates to 117,250 (median, 95% CrI: 44,480-447,800) averted symptomatic infections across the range of clinical severity. However, based on our representative surveys, only 1 in 50 (mean: 2.0%, 95% CI: 1.1%–3.5%) people with acute watery diarrhea, across the spectrum of severity, would have sought care in one of the cholera treatment centers in this study. We therefore estimate that the OCV campaign averted 2,350 (median, 95% CrI: 890-8,960) true cholera cases that would have been captured by the surveillance system in the three years post-vaccination, of which 840 (95% CrI: 350-4,360) averted in the first, 250 (95% CrI: 120-740) in the second and 1,240 (95% CrI: 140-4,850) in the third year post-vaccination (Supplementary Table S6). We estimate that 44 (median, 95% CrI: 16-150) cholera deaths were averted by the vaccination campaigns, including 20 (median, 95% CrI: 7-67) that would have been recorded by the surveillance system.

**Figure 3.**
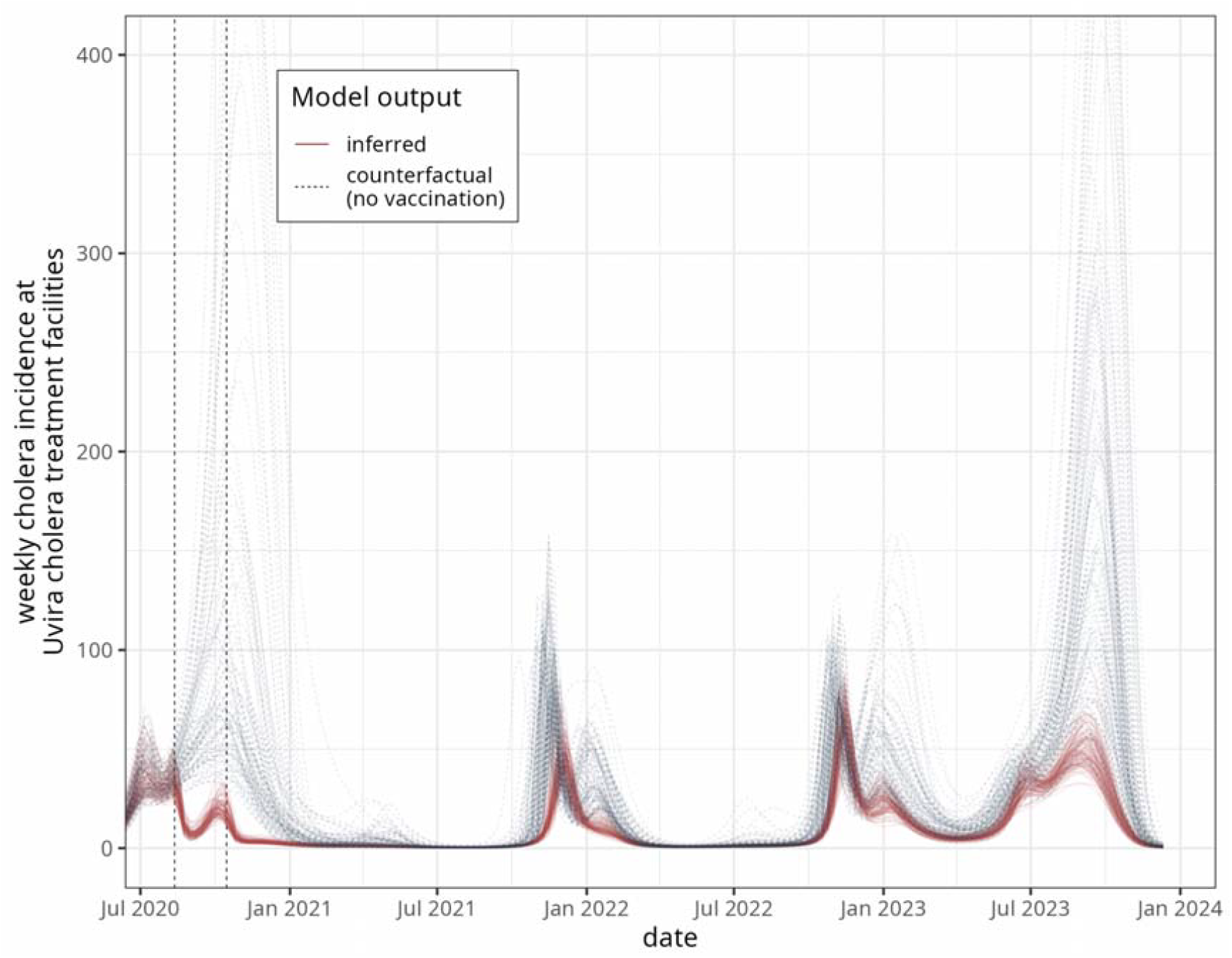
Counter-factual analysis of the impact of the OCV campaign in Uvira, DRC. Modeled weekly cholera incidence at Uvira cholera treatment facilities (red) is compared to a counterfactual scenario with no vaccination (navy blue, dashed line). Red lines indicate the mean and ribbons the 95% CrI of 2,000 posterior draws of inference for the stacked modeled outputs (see Methods). Blue lines indicate 100 draws from the counterfactual scenarios of the stacked modeled outputs. The y-axis was limited to 400 cases/week, with the maximum values across HMC draws of 1,036 cases/week in September 2023.

Sensitivity analyses show that our main results are robust to several key modeling assumptions. We find that impact was slightly lower (42%, 95% CrI: 31-63) when assuming health seeking increased after the implementation of our surveillance study in the CTCs in 2020. Impact was slightly larger (66%, 95% CrI: 32-88) if 200-day seroincidence in May 2022 is assumed to be 60% instead of 37.9% as in the main analysis, but lower (37%, 95% CrI: 26-53) when assuming only 15% seroincidence, although uncertainty intervals overlapped with the main estimates (Supplementary Table S6).

Effective vaccine coverage, which is determined by campaign coverage and population turnover, shapes vaccine impact. The ensemble models illustrate that OCV impact in Uvira does not have a linear relation with vaccination coverage, with diminishing impact efficiency above approximately 50% coverage. If the entire eligible population (all aged 1 year and above, 96.3% of the population^16^) had been vaccinated with at least one dose, 64% (median, 95% CrI: 43-87) of cholera cases and deaths would have been averted (Figure 4). The number of averted cases per vaccine dose had a decreasing trend from 2.1 (95% CrI: 0.5-10.3) cases-per-dose at 15% coverage to 0.5 (95% CrI: 0.2-1.7) at full eligible coverage (Supplementary Figure S6). We find that population turnover can diminish impact due to dilution of coverage, although with considerable uncertainty. If the population turnover would have been half of that observed in Uvira, the impact would have been slightly higher at 71% (median, 95% CrI: 52-90) of cases averted, while settings with doubled population turnover would have decreased the impact to 50% of cases averted (median, 95% CrI: 30-79).

**Figure 4.**
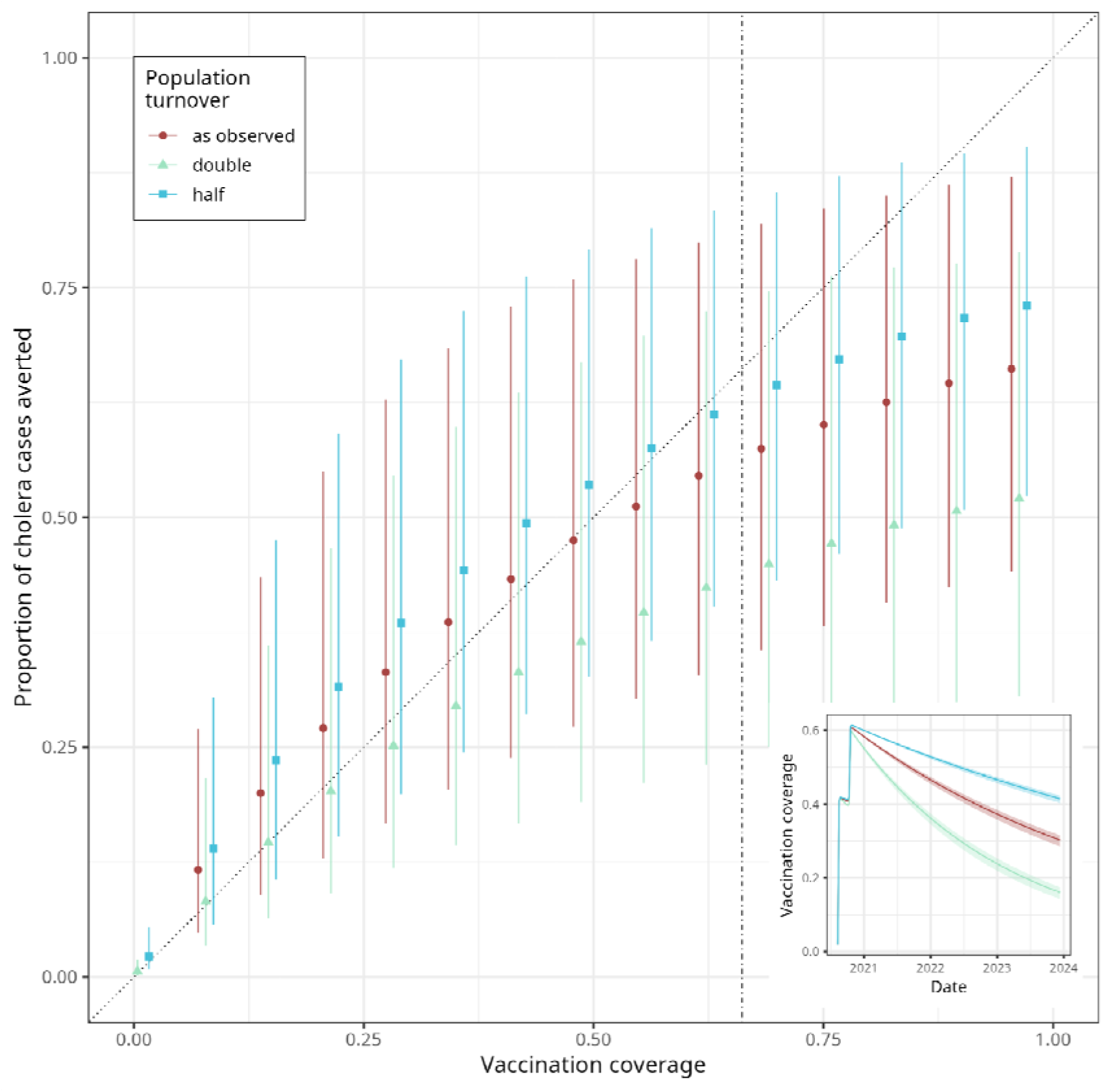
Counter-factual analysis of the effect of initial vaccination coverage on overall OCV impact in Uvira, DRC. Fraction of averted cases by the 2020 OCV campaign over a three-year evaluation period after campaign start as a function of initial vaccination coverage and population turnover scenarios. Population turnover scenarios include that which was inferred in the analysis (red dots), and additional scenarios with half (blue squares) and double (green triangles) the observed turnover. Points indicate the mean and bars the 95% credible intervals of 2,000 posterior draws of the ensemble modeled draws (see Methods). The vertical dot-dashed line indicates the initial vaccination coverage as inferred in this analysis. The inset figure shows changes in the effective vaccine coverage in time for the different population turnover scenarios.

## Discussion

Using detailed clinical, diagnostic, serologic, and demographic data, we estimated the three-year impact of the 2020 mass vaccination campaign in the cholera-endemic city of Uvira, eastern DRC, a long-standing research priority of the WHO and GTFCC.^3,11,12^ With 66% of the population receiving at least one OCV dose, the campaign averted a median of 56% of cases and deaths over three years. This corresponds to 117,250 averted symptomatic infections in the community, of which 2,350 would have sought care at the cholera treatment centers in Uvira. To our knowledge this is the first estimate of OCV impact in a cholera-endemic setting in Africa, falling in between previous randomized trial evaluations of overall two-dose protective effectiveness from Bangladesh (37%^9^) and India (75%^10^).

OCV impact in endemic settings may not be readily visible in surveillance data alone, as observed incidence may not decline markedly after vaccination, given variations in transmission intensity and imperfect vaccine protection and coverage. Contrary to expectations from previous modeling studies, the mass vaccination campaigns in Uvira did not lead to sustained suppression of transmission.^5^ By integrating uniquely rich epidemiological, serological and healthcare-seeking survey data, this study allowed us to infer transmission and immunity dynamics, thus unlocking counterfactual modeling for impact estimation. Notably, with our model structures, immune protection strength and duration were found to differ between symptomatic (strong, on the scale of years) and asymptomatic (weaker, on the scale of weeks), echoing modeling results based on historical data from the former British East Indian province of Bengal, although most other model conceptualizations assume a single unified post-infection immunologic profile.^23^

The transmission intensity we infer in Uvira has practical implications for cholera control in endemic settings. Our mean R_0_ estimates in the range of 10 are, to our knowledge, the first that are serology-informed, and far exceed the range of 1.5 to 3.5 in most publications, though models accounting for a hyper-infectious transient bacterial state have defended values of this magnitude.^29^ This discrepancy likely reflects earlier assumptions about relatively low population immunity,^30,31^ in contrast to a recent analysis from eastern DRC also finding high immunity and R_0_ values.^31^ Changes in transmissibility may be driven by flooding ^32^, WASH interventions during the study period,^33^ or COVID-19-era behavior changes. If our estimates of high transmission match the reality in Uvira and only a fraction of infections conferring durable immunity, multisectoral approaches rather than mass vaccination alone are required for sustained control. The large seasonal fluctuations in the reproductive number estimates further suggest that seasonally-timed WASH, OCV, or prophylactic antibiotics interventions could have an important impact.^34^

Our results offer several lessons for future preventive OCV campaigns in high-endemicity settings. First, the proportion of cases averted depends on background transmission. Lower infection rates in India than in Bangladesh, where annual sero-incidence estimates can exceed 50%, may in part explain the gap between existing protection estimates.^28,35^ Second, vaccine impact does not scale linearly with local vaccination coverage.^36^ Increasing at least one-dose coverage by 30 percentage points to reach the entire eligible population would have raised median impact by only 8 percentage points, as averted-cases-per-dose declines with coverage. Third, vaccination impact may be limited by coverage dilution due to population turnover, which we estimate in Uvira at a rate of 18% per year.^16^ This reinforces evidence from elsewhere in DRC that the spatial scale of campaigns shape impact in patchily vaccinated areas with high mobility.^36^ It further suggests that rather than pursuing 90-95% coverage targets in specific high-burden communities, more modest coverage across areas that account for the geographic scale of migration may deliver comparable population-level impact for substantially fewer resources.^36^ Finally, evaluation time frames must account for transmission and immunity dynamics, as impact fell from 79% of cases in the first year to around 45% thereafter, illustrating both how impact may vary in time, but also how vaccination can avert cases across cholera seasons in endemic settings.

Our study estimated OCV impact accounting for multiple sources data and uncertainty, but comes with limitations. We did not differentiate one-from two-protection or protection by age, following prior evidence from Uvira during the study period. Since young children make up roughly 15% of the population, substantially lower protection in this group would only slightly reduce our estimate, depending on mixing patterns,^17^ and our one-dose protection estimates were similar to published two-dose values.^6,16,17^ While we considered a large suit of different models, our inference on both impact and natural history of cholera is conditional on the assumptions within each. For example, additional model complexity, including contact network structure and mixing by risk group could lead to differing results though we have insufficient data to parameterize such complex models. Health-seeking behavior linking modeled infections to observed facility cases was measured by surveys and may be subject to desirability bias or fail to represent the pre-vaccination period, although sensitivity analysis showed that our estimates were robust to realistic variations in healthcare seeking and plausible biases in seroincidence. Finally, we may be underestimating the overall impact of vaccination, as genomic and epidemiologic evidence indicates high connectivity between Uvira’s South Kivu province and nearby regions inside and outside of DRC,^37^ so reducing incidence in Uvira may also have limited outbreak seeding elsewhere.

Real-world evidence of vaccine impact is increasingly important as global funding contracts and vaccine supply remains constrained. Our findings show that mass OCV campaigns can avert a substantial share of cases and deaths even in a hyperendemic setting where outbreaks continue and these benefits may not be apparent from surveillance trends alone. At the same time, the resurgence of cholera in Uvira within two years of a moderate-coverage campaign is a clear reminder that vaccination, however well delivered, cannot by itself interrupt transmission where the force of infection remains high. These findings should not be interpreted as evidence that OCV has failed, but rather that their substantial benefits can coexist with continued transmission. Durable cholera control will require sustained investment in safe water and sanitation, surveillance and healthcare system strengthening, with OCV as one component of an integrated strategy. Continued measurement of vaccine impact will be essential to ensure that gains from vaccination are neither overlooked nor overstated, and that expectations for what OCV can achieve remain realistic and evidence-based.

## Data Availability

Data and code needed to reproduce primary analyses are available at https://github.com/GenevaIDD/uvira-ocv-impact.

## Acknowledgements

The study team would like to thank Joël Faraja Zigashane Mashauri, Faraja Masembe Lulela, Jean-Marie Masugamuhanya Cirhonda and all the field teams in Uvira who have contributed to the cholera research program, including staff working in the surveys and clinical aspects of the study. The Uvira Health Zone, led by Dr. Panzu Nimi, played an important role in helping us to work smoothly with the local health system and communicate with the community. The study team thanks Maya Demby for her support throughout the entire program and Madiha Shafquat on some early efforts to model cholera in Uvira. The team would like to thank the Steering Committee who provided valuable feedback to help steer the course of activities throughout the study. Finally, the team would like to thank all participants in all aspects of our research program.

## Author Contributions

Conceptualization: EBM, KG, DM, ASA; Data curation: JJ, ET, JDH; Formal Analysis: JP, AD; Funding acquisition: KG, JKn, ASA; Investigation: EBM, PMB, CH, ET, BBR, MI, CC, DTL; Methodology: JP, JAB, CH, ASA; Project administration: EBM, PMB, JKn, ASA; Resources:; Software: JP, JAB; Supervision: JP, EBM, CH, DTL, KG, DM, JKn, ASA; Validation: JP, EBM, JJ, JDH, JKn, ASA; Visualization: JP; Writing – original draft: JP, EBM, ASA; Writing – review & editing: JP, EBM, JAB, PMB, CH, JJ, ET, JDH, JMS, BBR, MI, OC, EG, JK, AD, JL, KG, ECL, PWO, DM, JKn, ASA

## Notes

### Competing Interest Statement

The authors have declared no competing interest.

### Author Declarations

The study was approved by the ethics committee at the Johns Hopkins Bloomberg School of Public Health (IRB00015785), the London School of Hygiene and Tropical Medicine (25365) and the University of Kinshasa School of Public Health (ESP/ CE/65/2021).

## References

1 Izawa Y, Ikejezie J, Escobar Corado Waeber RA, et al. Global resurgence of cholera, 2022–2025: Epidemiological insights from WHO surveillance data. Social Science Research Network. 2026; published online March 18. DOI:10.2139/ssrn.6431513.

2 Local Burden of Disease WaSH Collaborators. Mapping geographical inequalities in access to drinking water and sanitation facilities in low-income and middle-income countries, 2000-17. Lancet Glob Health 2020; 8: e1162–85.

3 WHO | Ending Cholera. 2017; published online Oct 31. http://www.who.int/cholera/publications/global-roadmap/en/ (accessed Nov 1, 2017).

4 Burki T. The great cholera vaccine shortage. Lancet 2024; 403: 891–2.

5 Longini IM Jr, Nizam A, Ali M, Yunus M, Shenvi N, Clemens JD. Controlling endemic cholera with oral vaccines. PLoS Med 2007; 4: e336.

6 Xu H, Tiffany A, Luquero FJ, et al. Protection from killed whole-cell cholera vaccines: a systematic review and meta-analysis. Lancet Glob Health 2025; 13: e1203–12.

7 Peak CM, Reilly AL, Azman AS, Buckee CO. Prolonging herd immunity to cholera via vaccination: Accounting for human mobility and waning vaccine effects. PLoS Negl Trop Dis 2018; 12: e0006257.

8 Ali M, Sur D, You YA, et al. Herd protection by a bivalent killed whole-cell oral cholera vaccine in the slums of Kolkata, India. Clin Infect Dis 2013; 56: 1123–31.

9 Qadri F, Ali M, Chowdhury F, et al. Feasibility and effectiveness of oral cholera vaccine in an urban endemic setting in Bangladesh: a cluster randomised open-label trial. Lancet 2015; 386: 1362–71.

10 Ali M, Debes AK, Luquero FJ, et al. Potential for controlling cholera using a ring vaccination strategy: Re-analysis of data from a cluster-randomized clinical trial. PLoS Med 2016; 13: e1002120.

11 Weekly epidemiological record Relevé épidémiologique hebdomadaire. 2017. https://iris.who.int/server/api/core/bitstreams/1c1a256a-e188-45b3-9987-0055fcf0235b/content (accessed Nov 25, 2025).

12 Cholera Roadmap Research Agenda. https://www.gtfcc.org/resources/cholera-roadmap-research-agenda/ (accessed Nov 25, 2025).

13 Azman AS, Rumunu J, Abubakar A, et al. Population-Level Effect of Cholera Vaccine on Displaced Populations, South Sudan, 2014. Emerg Infect Dis 2016; 22: 1067–70.

14 Schyns C, Fossa A, Mutombo-Nfenda, et al. Cholera in Eastern Zaire, 1978. Ann Soc Belg Med Trop 1979; 59: 391–400.

15 Jeandron A, Cumming O, Rumedeka BB, Saidi JM, Cousens S. Confirmation of cholera by rapid diagnostic test amongst patients admitted to the cholera treatment centre in Uvira, Democratic Republic of the Congo. PLoS One 2018; 13. DOI:10.1371/journal.pone.0201306.

16 Koyuncu A, Bugeme PM, Dent J, et al. Challenges with achieving and maintaining oral cholera vaccine coverage: insights from serial cross-sectional representative surveys in a cholera-endemic community in the Democratic Republic of the Congo. BMJ Public Health 2025; 3: e001035.

17 Malembaka EB, Bugeme PM, Hutchins C, et al. Effectiveness of one dose of killed oral cholera vaccine in an endemic community in the Democratic Republic of the Congo: a matched case-control study. Lancet Infect Dis 2024; 24: 514–22.

18 Jones FK, Bhuiyan TR, Muise RE, et al. Identifying recent cholera infections using a multiplex bead serological assay. MBio 2022; 13. DOI:10.1128/mbio.01900-22.

19 Rogan WJ, Gladen B. Estimating prevalence from the results of a screening test. Am J Epidemiol 1978; 107: 71–6.

20 Jones FK, Bhuiyan TR, Slater DM, et al. Expanding cholera serosurveillance to vaccinated populations. mBio 2025; published online Nov 12. DOI:10.1128/mbio.01898-25.

21 Bouman JA, Hauser A, Grimm SL, et al. Bayesian workflow for time-varying transmission in stratified compartmental infectious disease transmission models. PLoS Comput Biol 2024; 20: e1011575.

22 Le A, King AA, Magpantay FMG, Mesbahi A, Rohani P. The impact of infection-derived immunity on disease dynamics. J Math Biol 2021; 83: 61.

23 King AA, Ionides EL, Pascual M, Bouma MJ. Inapparent infections and cholera dynamics. Nature 2008; 454: 877–80.

24 Muzembo BA, Kitahara K, Debnath A, Okamoto K, Miyoshi S-I. Accuracy of cholera rapid diagnostic tests: a systematic review and meta-analysis. Clin Microbiol Infect 2022; 28: 155–62.

25 Vehtari A, Gabry J. Bayesian Stacking and Pseudo-BMA weights using the loo package. Version loo 2019. https://ftp.sun.ac.za/ftp/CRAN/web/packages/loo/vignettes/loo2-weights.html.

26 Yao Y, Vehtari A, Simpson D, Gelman A. Using stacking to average Bayesian predictive distributions (with discussion). Bayesian Anal 2018; 13: 917–1007.

27 Bugeme PM, Xu H, Hutchins C, et al. Cholera Deaths During Outbreaks in Uvira, Eastern Democratic Republic of the Congo, 10-35 Months After Mass Vaccination. Open Forum Infect Dis 2024; 11: ofae058.

28 Hegde S, Khan A, Perez-Saez J, et al. Clinical surveillance systems obscure the true cholera infection burden in an endemic region. Nature Medicine 2024; 30: 888–95.

29 Hartley DM, Morris JG Jr, Smith DL. Hyperinfectivity: a critical element in the ability of V. cholerae to cause epidemics? PLoS Med 2006; 3: e7.

30 Chan CH, Tuite AR, Fisman DN. Historical epidemiology of the second cholera pandemic: relevance to present day disease dynamics. PLoS One 2013; 8: e72498.

31 Blake A, Walder A, Hanks EM, et al. Impact of a multi-pronged cholera intervention in an endemic setting. PLOS Neglected Tropical Diseases 2025; 19: e0012867.

32 Alcayna T, Fletcher I, Gibb R, et al. Climate-sensitive disease outbreaks in the aftermath of extreme climatic events: A scoping review. One Earth 2022; 5: 336–50.

33 Gallandat K, Macdougall A, Jeandron A, et al. Improved water supply infrastructure to reduce acute diarrhoeal diseases and cholera in Uvira, Democratic Republic of the Congo: Results and lessons learned from a pragmatic trial. PLoS Negl Trop Dis 2024; 18: e0012265.

34 Perez-Saez J, Lessler J, Lee EC, et al. The seasonality of cholera in sub-Saharan Africa: a statistical modelling study. The Lancet Global Health 2022; 10: e831–9.

35 Azman AS, Lauer SA, Bhuiyan TR, et al. Vibrio cholerae O1 transmission in Bangladesh: insights from a nationally representative serosurvey. Lancet Microbe 2020; 1: e336–43.

36 Briskin E, Bateyi Mustafa SH, Mahamba R, et al. Oral cholera vaccine coverage in Goma, Democratic Republic of the Congo, 2022, following 2019-2020 targeted preventative mass campaigns. Vaccine X 2024; 20: 100555.

37 DiPrete BL, Perez-Saez J, Wohl S, et al. Defining Epidemiologically Relevant Units of Cholera Transmission in sub-Saharan Africa. https://www.medrxiv.org/content/10.1101/2025.06.06.25329161v1.

