## Supplementary file for "Impact of mass oral cholera vaccination in an endemic area of the Democratic Republic of the Congo: a surveillance-based counterfactual modeling analysis"

#### Contents

**S1 Luminex assay** 2

**S2 Wavelet analysis** 3

**S3 Vaccine effectiveness estimation with the screening method** 3

**S4 Modeling analyses** 3

### S1 Luminex assay

We used the Luminex MAGPIX to assess antibody levels in serum collected from our representative serosurveys in Uvira. We conjugated fluorescent magnetic microspheres (MagPlex-C Microspheres, Luminex MC100 series) to their corresponding antigens using the xMAP Antibody Coupling Kit (Luminex cat# 40-50016) according to the manufacturer's instructions. Antigen-conjugated magnetic microspheres (beads) were counted and performance was titrated to 1200 beads per antigen per well. The antigens used were sourced and conjugated to specific bead regions as follows: *V. cholerae* Inaba OSP-BSA (donation from Dr. Edward Ryan, region 44), *V. cholerae* Ogawa OSP-BSA (donation from Dr. Edward Ryan, region 42), cholera toxin subunit B (CtxB, Sigma C9903-1MG, region 38). We used an eight-point, 4-fold dilution curve of pooled convalescent plasma from culture-confirmed cholera patients to estimate relative antibody concentrations, beginning at a dilution of 1:100 and ending at a dilution of 1:1638400. Four full standard curves were run in duplicate throughout the course of the lab analyses to account for variation over time. Each assay plate included high and low concentrations of standard for each antigen (1:100 and 1:1000 respectively), run in duplicate, which were used for quality control. Samples were tested with a final dilution of 1:1000. All sample and control dilutions were performed using phosphate buffered saline (PBS) with 1% bovine serum albumin (BSA) and 0.05% Tween-20 (sample dilution buffer). Each plate also contained two blank wells of sample dilution buffer to account for background fluorescence. Plates with a high background or samples with a bead count of less than 30 were re-run. Samples were pre-diluted and aliquoted within the week and stored sealed at 4°C until use.

For the assay, all reagents were allowed to equilibrate to room temperature for 1 hour, and a master mix of beads for each plate was prepared. Antigen-conjugated magnetic beads were incubated with sample or control sera at room temperature for 1.5 hours, covered with foil and shaking vigorously at 700-900 RPM. Following incubation with sera, beads were washed by magnetic separation three times in PBS with 0.05% Tween-20 and incubated with biotinylated anti-human IgG secondary antibody (50 ng per well) for 45 minutes at room temperature, shaking as described at 700-900 RPM. Following another series of washes as described above, beads were incubated with streptavidin conjugated to phycoerythrin (SA-PE, Invitrogen cat# S866, 250 ng per well), shaking for 30 minutes. Samples were washed and incubated with PBS with 0.5% BSA and 0.05% Tween-20 as an extended wash step for 30 minutes shaking at 700-900 RPM. Beads were then washed by magnetic separation a final time before resuspending in PBS. We stored samples overnight at 4°C before analyses on the MAGPIX instrument. Prior to each instrument run, the MAGPIX passed an instrument quality control verification, and a calibration test at least weekly (ThermoFisher MPXPVERK25 and MPXCALK25 respectively). Due to instrument performance issues, two different MAGPIX instruments were used through the course of the study. Standard curves were run and compared between instruments.

| **Antigen** | **Source** | **Beads per well** | **Bead Region** | **Standard** | **4-Fold Standard Curve Starting Point** |
|---|---|---|---|---|---|
| *Vibrio cholerae* Inaba OSP-BSA | Edward Ryan | 1200 | 44 | Cholera convalescent plasma (MGH/Hopkins) | 1:100 |
| *Vibrio cholerae* Ogawa OSP-BSA | Edward Ryan | 1200 | 42 | Cholera convalescent plasma (MGH/Hopkins) | 1:100 |
| Cholera toxin subunit B (CtxB) | Sigma C9903-1 MG | 1200 | 38 | Cholera convalescent plasma (MGH/Hopkins) | 1:100 |

Table S1: Luminex antigen overview.

#### S2 Wavelet analysis

To characterize the periodicity of cholera transmission over the surveillance period, we performed a continuous wavelet transform of the weekly cholera incidence series using the Morlet wavelet (R package WaveletComp version 1.2). We reconstructed a proxy of weekly cholera incidence that accounts for the fact that laboratory confirmation was only available in a subset of suspected cases. To do this, we first modeled a weekly positivity rate with a binomial GAM, fitting a spline of time to weekly RDT-positive and RDT-test counts. Then we multiplied the smoothed weekly positivity rate and the weekly number of suspected AWD cases to generate the weekly cholera incidence proxy. To characterize the periodicity of cholera incidence we then applied the wavelet transform to the square root of this reconstructed series at weekly resolution and over periods ranging from 4 to 255 weeks (˜5 years). The significance of the wavelet power was assessed against a white-noise null distribution by simulation as implemented in WaveletComp.

#### S3 Vaccine effectiveness estimation with the screening method

As an independent check on the vaccine effectiveness (VE) implied by the transmission model, we also estimated the effectiveness of at least one dose of OCV directly from the surveillance data using the screening method, which contrasts the proportion of cases who had been vaccinated with the vaccine coverage in the source population [1].

**Population vaccine coverage.** We estimated time-varying coverage $c(t)$ from the population-representative coverage surveys by fitting a logistic regression of individual vaccination status on the number of weeks since the first vaccination round, $w$,

$$\operatorname{logit} c(w) = \alpha + \beta\, w,$$

and predicting coverage for each week of the post-campaign period.

**Proportion of cases vaccinated.** For each calendar month $t$ of the post-campaign period we computed the proportion of confirmed (RDT-positive) cholera cases who reported prior vaccination,

$$\mathrm{PCV}(t) = \frac{y_v(t)}{y(t)},$$

where $y(t)$ is the number of RDT-positive cases with known vaccination status and $y_v(t)$ the subset who were vaccinated with at least one dose of OCV.

**Effectiveness.** Under the screening method, VE is one minus the odds ratio of vaccination among cases relative to the population,

$$\mathrm{VE}(t) = 1 - \frac{\mathrm{PCV}(t)/\left(1 - \mathrm{PCV}(t)\right)}{c(t)/\left(1 - c(t)\right)}.$$

Point estimates and 95% confidence intervals were obtained by fitting, for each month, an intercept-only binomial regression for the vaccinated case count $y_v(t)$ out of $y(t)$ with the logit of the estimated population coverage as a fixed offset,

$$y_v(t) \sim \mathrm{Binomial}\big(y(t),\, p(t)\big), \qquad \operatorname{logit} p(t) = \theta(t) + \operatorname{logit} c(t),$$

so that $\mathrm{VE}(t) = 1 - \operatorname{logit}^{-1}\big(\hat{\theta}(t)\big)$, with the confidence interval following from the Wald interval for $\theta(t)$. Months with no RDT-positive cases were omitted. These screening-method estimates are shown alongside the model-implied effectiveness in panel c of Figure S4.

#### S4 Modeling analyses

The primary aim of this modeling analysis was to estimate the impact in terms of averted cholera cases and deaths, of the 2020 OCV campaign in Uvira, DRC. The secondary aim of the model was to infer natural history and immune dynamics parameters of cholera in this endemic setting. To this end we developed a compartmental SIAR-type model framework for cholera transmission to leverage the different data streams of this study which included epidemiologic surveillance as well as population-representative vaccine coverage and serological surveys. We then use this modeling framework in counterfactual analyses to estimate the impact of the 2020 OCV campaign under a set of alternative scenarios of vaccine coverage and population turnover.

Code is available at https://github.com/GenevaIDD/uvira-ocv-impact.

#### S4.1 Transmission model description

##### S4.1.1 Model formulations

We develop a suite of compartmental models of cholera transmission in Uvira, DRC, building on published cholera models that follow the SIAR-type framework [2, 3], which have also been used to assess the impact of OCV [4]. Given the primary aim of the analysis and available data streams, key components of the model to account for were: i) OCV-induced immunity, ii) population turnover in Uvira leading to decline in vaccine coverage, iii) asymptomatic infections to coordinate modeled outputs to serological data (see Section **??**).

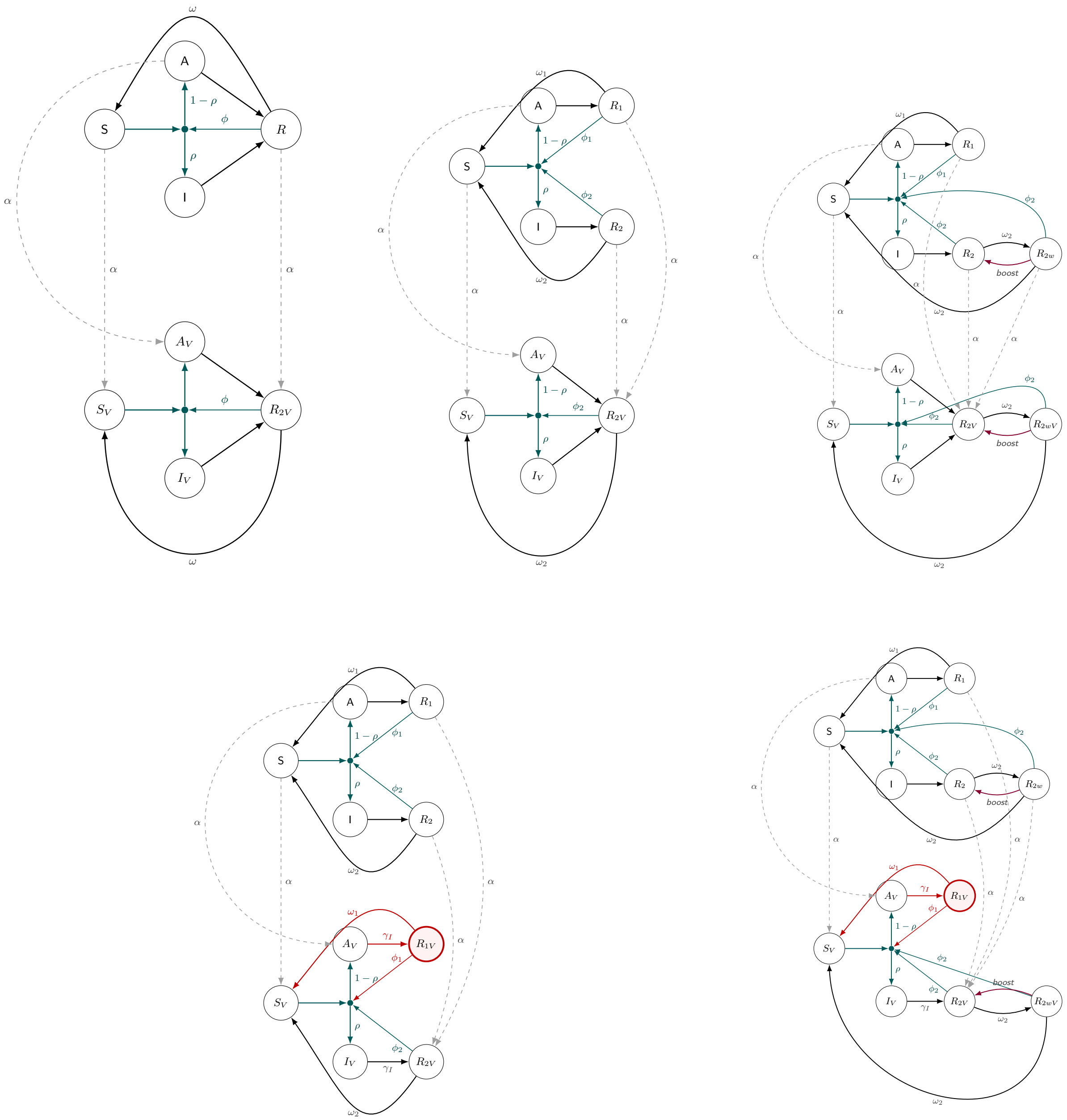

Figure S1: Model diagrams. Top row, left to right: SIAR (root), two-path, and two-path-boost model formulations. Bottom row: the corresponding variants with mirroring of two-path immunity in the vaccinated compartments (new compartments and flows in red). In each panel the upper block is the unvaccinated arm and the lower block the vaccinated arm. Teal: infection. Susceptibles and all partially protected compartments feed a single junction (filled dot) representing the force of infection $\lambda(t)$, which then splits into symptomatic infection with probability $\rho$ and asymptomatic infection with probability $1-\rho$; the thin teal arrows entering the junction carry the leaky-protection parameter ($\phi$, $\phi_1$ or $\phi_2$), so that individuals in these compartments, including vaccinated ones, are (re)infected at the reduced rate $\phi\lambda(t)$. Black: recovery (rate $\gamma_I$) and waning of immunity (rates $\omega$, or $\omega_1$ and $\omega_2$). Purple: immune boosting on re-exposure ($R_{2w} \to R_2$, $R_{2wV} \to R_{2V}$). Dashed grey: vaccination at rate $\alpha(t)$. Demographic flows (births, immigration, emigration, cholera mortality), which apply to all compartments, are omitted for clarity. Compartment definitions, equations and parameters are given in the text and in Table S3; structural differences between formulations are summarised in Table S2.

**S4.1.1.1 SIAR model** We first consider an SIR model with an asymptomatic compartment. In this first model susceptible individuals (S) can become infected through contacts with infected and symptomatic individuals (I) or asymptomatic individuals (A), both leading to a resistant/immune compartment (R). Infections are symptomatic with probability $\rho$, and infectiousness lasts an average of $1/\gamma_I$. We here model cholera immunity using a leaky-immunity model where infections provides imperfect initial protection ($\phi \in [0,1]$) which further wanes with time post-infection with individuals returning to a fully susceptible state at an exponential rate $\omega$ [5]. We note that here symptomatic and asymptomatic infections are assumed to lead to the same immune state. The total population in Uvira may change through births (at rate $\mu_B$), emigration (at rate $\mu_{\mathcal{E}}$) and immigration (at rate $\mu_{\mathcal{I}}$), with the assumption that the composition of the immigrating population is the same as the one in Uvira at the time of immigration. Vaccination is represented by a vaccinated compartment ($R_{2V}$) assumed to have the same immune protection properties as R, with waning of immunity leading to a susceptible-and-vaccinated compartment ($S_V$) which has the same properties as S but is kept separate for accounting purposes to allow for model coordination with epidemiological data. The SIAR model is defined as follows:

$$
\begin{aligned}
\frac{dS}{dt} &= -\lambda(t)S + \omega R + (\mu_B + \mu_{\mathcal{I}} f_S(t))N - (\mu_{\mathcal{E}} + \alpha(t))S, \\
\frac{dI}{dt} &= \rho\lambda(t)(S + \phi R) - (\gamma_I + \mu_I)I + \mu_{\mathcal{I}} f_I(t)N, \\
\frac{dA}{dt} &= (1-\rho)\lambda(t)(S + \phi R) - (\gamma_I + \mu_{\mathcal{E}} + \alpha(t))A + (\mu_{\mathcal{I}} f_A(t))N, \\
\frac{dR}{dt} &= \gamma_I(I + A) - (\lambda(t)\phi + \omega + \mu_{\mathcal{E}} + \alpha(t))R + \mu_{\mathcal{I}} f_R(t)N, \\
\frac{dS_V}{dt} &= -\lambda(t)S_V + \omega R_{2V} - \mu_{\mathcal{E}} S_V, \\
\frac{dI_V}{dt} &= \rho\lambda(t)(S_V + \phi R_{2V}) - (\gamma_I + \mu_I + \mu_{\mathcal{E}})I_V, \\
\frac{dA_V}{dt} &= \alpha(t)A + (1-\rho)\lambda(t)(S_V + \phi R_{2V}) - (\gamma_I + \mu_{\mathcal{E}})A_V, \\
\frac{dR_{2V}}{dt} &= \alpha(t)(S + R) + \gamma_I(I_V + A_V) - (\lambda(t)\phi + \omega + \mu_{\mathcal{E}})R_{2V},
\end{aligned}
$$

where $\lambda(t)$ is the time-varying force of infection, $N = S + I + A + R + S_V + I_V + A_V + R_{2V}$ is the total population, $\alpha(t)$ is the vaccination rate (see below) and $f_S, f_I, f_A, f_R$ are the fractions of each compartment S, I, A and R in the total unvaccinated population: $f_X = X/(S + I + A + R)$, with the exception of I $f_I = (X + \xi)/(S + I + A + R)$, where $\xi$ represents a small influx of infected individuals. Model parameters definitions and values are given in table S3.

We model the time-varying force of infection as:

$$
\lambda(t) = \beta(t)\left(I + I_V + \beta_A(A + A_V)\right),
$$

where $\beta(t)$ is a time-varying transmission rate, and $\beta_A \in [0,1]$ represents the reduced infectiousness of those with an asymptomatic infection.

**S4.1.1.2 Basic reproduction number** For inference purposes we reparametrize the transmission rate in terms of the time-varying basic reproduction number $R_0(t)$ and the other model parameters. We derive $R_0$ using the next generation matrix method [6]. At the disease-free equilibrium the whole (normalized) population is susceptible ($S = N$, all other compartments empty), and in the absence of vaccination ($\alpha(t) = 0$) the only infected states are the symptomatic ($I$) and asymptomatic ($A$) compartments. Linearizing the inflow of new infections and the transitions of the infected subsystem $(I, A)$ about this equilibrium gives the new-infection matrix $\mathbf{F}$ and the transition matrix $\mathbf{V}$:

$$\mathbf{F} = \beta(t) \begin{pmatrix} \rho & \rho\,\beta_A \\ 1-\rho & (1-\rho)\,\beta_A \end{pmatrix}, \qquad \mathbf{V} = \begin{pmatrix} \gamma_I + \mu_I & 0 \\ 0 & \gamma_I + \mu_{\mathcal{E}} \end{pmatrix},$$

where the columns of $\mathbf{F}$ give the rates at which the symptomatic and asymptomatic classes generate new infections (a fraction $\rho$ symptomatic and $1-\rho$ asymptomatic, with asymptomatic carriers contributing a factor $\beta_A$), and $\mathbf{V}$ collects the exit rates from each infected class: recovery $\gamma_I$ together with cholera mortality $\mu_I$ for the symptomatic class and emigration $\mu_{\mathcal{E}}$ for the asymptomatic class. The basic reproduction number is the spectral radius of the next generation matrix $\mathbf{K} = \mathbf{F}\,\mathbf{V}^{-1}$. As $\mathbf{F}$ has rank one, the spectral radius equals the trace of $\mathbf{K}$, giving:

$$R_0(t) = \beta(t) \left( \frac{\rho}{\gamma_I + \mu_I} + \frac{(1-\rho)\,\beta_A}{\gamma_I + \mu_{\mathcal{E}}} \right).$$

Because the infected subsystem $(I, A)$ and the structure of new infections are identical across all model formulations considered here — the variants differ only in their recovery and vaccinated compartments, which are empty at the disease-free equilibrium — this expression for $R_0$ is the same for every model.

Assuming no population turnover ($\mu_I = \mu_{\mathcal{E}} = 0$), this simplifies to

$$R_0(t) = \frac{\beta(t)\,(\rho + (1-\rho)\beta_A)}{\gamma_I},$$

and thus $\beta(t) = R_0(t)\gamma_I/(\rho + (1-\rho)\beta_A)$, the form used to reparametrize the transmission rate.

The no-turnover form used for the reparametrization differs from the full expression by the factor

$$\frac{R_0(t)}{R_0^{\text{turnover}}(t)} = \frac{\rho + (1-\rho)\beta_A}{\rho\,\dfrac{\gamma_I}{\gamma_I + \mu_I} + (1-\rho)\beta_A\,\dfrac{\gamma_I}{\gamma_I + \mu_{\mathcal{E}}}},$$

a weighted average (with weights $\rho$ and $(1-\rho)\beta_A$) of the shrinkage factors $\gamma_I/(\gamma_I+\mu_I) \le 1$ and $\gamma_I/(\gamma_I+\mu_{\mathcal{E}}) \le 1$, so that the no-turnover value never underestimates the turnover-adjusted one. Because we assume no symptomatic-infection mortality ($\mu_I = 0$) and the mean infectious period ($1/\gamma_I = 2$ days) is much shorter than the demographic timescale of emigration, both factors are close to one and the relative correction $\approx \frac{(1-\rho)\beta_A}{\rho+(1-\rho)\beta_A}\,\frac{\mu_{\mathcal{E}}}{\gamma_I}$ is well below 0.1%, negligible relative to the posterior credible interval on $R_0$.

We then modeled $R_0$ using basis-splines following Bouman et al. [7], with cubic splines and knots at 10-week intervals:

$$\log(R_0(t)) = \sum_{i=1}^{K} a_i B_{i,4}(t),$$

where $K$ is the total number of B-splines bases, $a_i$ are the spline coefficients and $B_{i,4}$ are the B-splines of order 4 (cubic B-splines).

The 2020 OCV campaign consisted of two separate rounds, the first in July and the second in October, we therefore model the vaccination rate as [8]:

$$\alpha(t) = \begin{cases} p\alpha & \text{if} \quad t \in (\text{July}, \text{July}), \\ (1-p)\alpha & \text{if} \quad t \in (\text{Oct}, \text{Oct}), \\ 0 & \text{else} \end{cases}$$

where $\alpha$ is a rate parameter to be inferred, and $p$ is the probability of having had the first vaccine dose in the first campaign round. We informed this probability using our 2021 representative household survey, where $p = 0.56$.

**S4.1.1.3 Two-path immunity** We extend the SIAR model to account for two distinct immunity compartments following King et al. [2]. In this model we differentiate immunity from asymptomatic ($R_1$) and from symptomatic infection ($R_2$), which are allowed to have different leaky immunity and immune waning parameters. Here vaccination is assumed to be equivalent to symptomatic infections ($R_2$) in terms of immune protection. The two-path immunity model then reads:

$$
\begin{aligned}
\frac{dS}{dt} &= -\lambda(t)S + \omega_1 R_1 + \omega_2 R_2 + (\mu_B + \mu_{\mathcal{I}} f_S(t))N - (\mu_{\mathcal{E}} + \alpha(t))S, \\
\frac{dI}{dt} &= \rho\lambda(t)(S + \phi_1 R_1 + \phi_2 R_2) - (\gamma_I + \mu_I)I + \mu_{\mathcal{I}} f_I(t)N, \\
\frac{dA}{dt} &= (1-\rho)\lambda(t)(S + \phi_1 R_1 + \phi_2 R_2) - (\gamma_I + \mu_{\mathcal{E}} + \alpha(t))A + (\mu_{\mathcal{I}} f_A(t))N, \\
\frac{dR_1}{dt} &= \gamma_I A - (\lambda(t)\phi_1 + \omega_1 + \mu_{\mathcal{E}} + \alpha(t))R_1 + \mu_{\mathcal{I}} f_{R_1}(t)N, \\
\frac{dR_2}{dt} &= \gamma_I I - (\lambda(t)\phi_2 + \omega_2 + \mu_{\mathcal{E}} + \alpha(t))R_2 + \mu_{\mathcal{I}} f_{R_2}(t)N, \\
\frac{dS_V}{dt} &= -\lambda(t)(S_V + \phi_2 R_{2V}) + \omega_2 R_{2V} - \mu_{\mathcal{E}} S_V, \\
\frac{dI_V}{dt} &= \rho\lambda(t)(S_V + \phi_2 R_{2V}) - (\gamma_I + \mu_I + \mu_{\mathcal{E}})I_V, \\
\frac{dA_V}{dt} &= \alpha(t)A + (1-\rho)\lambda(t)(S_V + \phi_2 R_{2V}) - (\gamma_I + \mu_{\mathcal{E}})A_V, \\
\frac{dR_{2V}}{dt} &= \alpha(t)(S + R_1 + R_2) + \gamma_I(I_V + A_V) - (\lambda(t)\phi_2 + \omega_2 + \mu_{\mathcal{E}})R_{2V}.
\end{aligned}
$$

**S4.1.1.4 Two-path immunity and boosting** Re-exposure to pathogens may lead to an immunological boost which can extend the duration of protection from natural infection or vaccination [5]. We therefore extend the two-path immunity model to account for boosting by introducing additional resistant and vaccinated compartments $R_{2w}, R_{2wV}$ to which individuals are re-assigned upon unsuccessful infections (i.e., boosting events):

$$
\begin{aligned}
\frac{dS}{dt} &= -\lambda(t)S + \omega_1 R_1 + \omega_2 R_{2w} + (\mu_B + \mu_{\mathcal{I}} f_S(t))N - (\mu_{\mathcal{E}} + \alpha(t))S, \\
\frac{dI}{dt} &= \rho\lambda(t)(S + \phi_1 R_1 + \phi_2(R_2 + R_{2w})) - (\gamma_I + \mu_I)I + \mu_{\mathcal{I}} f_I(t)N, \\
\frac{dA}{dt} &= (1-\rho)\lambda(t)(S + \phi_1 R_1 + \phi_2(R_2 + R_{2w})) - (\gamma_I + \mu_{\mathcal{E}} + \alpha(t))A + (\mu_{\mathcal{I}} f_A(t))N, \\
\frac{dR_1}{dt} &= \gamma_I A - (\lambda(t)\phi_1 + \omega_1 + \mu_{\mathcal{E}} + \alpha(t))R_1 + \mu_{\mathcal{I}} f_{R_1}(t)N, \\
\frac{dR_2}{dt} &= \gamma_I I + \lambda(t)(1-\phi_2)R_{2w} - (\lambda(t)\phi_2 + \omega_2 + \mu_{\mathcal{E}} + \alpha(t))R_2 + \mu_{\mathcal{I}} f_{R_2}(t)N, \\
\frac{dR_{2w}}{dt} &= \omega_2 R_2 - (\lambda(t)\phi_2 + \omega_2 + \mu_{\mathcal{E}} + \alpha(t))R_{2w} + \mu_{\mathcal{I}} f_{R_{2w}}(t)N, \\
\frac{dS_V}{dt} &= -\lambda(t)(S_V + \phi_2(R_{2V} + R_{2wV})) + \omega_2 R_{2wV} - \mu_{\mathcal{E}} S_V, \\
\frac{dI_V}{dt} &= \rho\lambda(t)(S_V + \phi_2(R_{2V} + R_{2wV})) - (\gamma_I + \mu_I + \mu_{\mathcal{E}})I_V, \\
\frac{dA_V}{dt} &= \alpha(t)A + (1-\rho)\lambda(t)(S_V + \phi_2(R_{2V} + R_{2wV})) - (\gamma_I + \mu_{\mathcal{E}})A_V, \\
\frac{dR_{2V}}{dt} &= \alpha(t)(S + R_1 + R_2 + R_{2w}) + \gamma_I(I_V + A_V) + \lambda(t)(1-\phi_2)R_{2wV} - (\lambda(t)\phi_2 + \omega_2 + \mu_{\mathcal{E}})R_{2V}, \\
\frac{dR_{2wV}}{dt} &= \omega_2 R_{2V} - (\lambda(t)\phi_2 + \omega_2 + \mu_{\mathcal{E}})R_{2wV}.
\end{aligned}
$$

Table S2: Structural features of the five candidate model formulations. Two-path immunity: separate immunity compartments for asymptomatic ($R_1$) and symptomatic ($R_2$) recovery with distinct leaky-protection parameters $\phi_1$, $\phi_2$ and waning rates $\omega_1$, $\omega_2$. Immune boosting: re-exposure events re-assign individuals from the waning compartment ($R_{2w}$) to a freshly boosted compartment, extending protection, with a free parameter $\nu$ scaling the $(1-\phi_2)$ re-exposure rate. Vaccinated asymptomatic recovery: an additional compartment $R_{1V}$ mirrors $R_1$ for vaccinated individuals recovering from asymptomatic infection, retaining the weaker $\phi_1$ protection instead of $\phi_2$. ✓: feature present.

| Model | Two-path immunity ($R_1/R_2$) | Immune boosting ($R_{2w}$, $\nu$) | Vaccinated asymptomatic recovery ($R_{1V}$) |
|---|---|---|---|
| SIAR | | | |
| two-path | ✓ | | |
| two-path-vacc-mirror | ✓ | | ✓ |
| two-path-boost | ✓ | ✓ | |
| two-path-boost-vacc-mirror | ✓ | ✓ | ✓ |

**S4.1.1.5 Vaccinated asymptomatic recovery** In the models above, vaccinated individuals recovering from asymptomatic infection ($A_V$) enter the vaccinated recovered compartment $R_{2V}$ and thereby acquire the stronger, symptomatic-type protection $\phi_2$. We additionally consider a set of variants in which these individuals instead enter a separate vaccinated asymptomatic-recovery compartment $R_{1V}$ that retains the weaker asymptomatic-type protection $\phi_1$ and wanes at rate $\omega_1$ to the vaccinated-susceptible compartment $S_V$, mirroring the $R_1$ compartment of the unvaccinated arm. Relative to the two-path model, the modified equations read:

$$\begin{aligned}
\frac{dR_{1V}}{dt} &= \gamma_I A_V - (\lambda(t)\phi_1 + \omega_1 + \mu_{\mathcal{E}})R_{1V}, \\
\frac{dR_{2V}}{dt} &= \alpha(t)(S + R_1 + R_2) + \gamma_I I_V - (\lambda(t)\phi_2 + \omega_2 + \mu_{\mathcal{E}})R_{2V}, \\
\frac{dS_V}{dt} &= -\lambda(t)(S_V + \phi_2 R_{2V}) + \omega_2 R_{2V} + \omega_1 R_{1V} - \mu_{\mathcal{E}} S_V,
\end{aligned}$$

with the remaining equations unchanged. The new compartment $R_{1V}$ contributes to the total population $N$, to the vaccinated population $N_V$, and to the effective susceptible population $X(t)$ with weight $\phi_1$. The same extension applies to the two-path-boost model. We call these sets of models "mirror" varriants.

#### S4.2 Population turnover

Uvira is subject to substantial population movement, and the resulting turnover of vaccinated individuals is an important driver of declining vaccine coverage. The ODE models include emigration and immigration parameters, for which we define priors based on available survey data.

We define our model priors based on data from our 2021–2023 representative household surveys, in which households (N=405) reported the number of members who had left Uvira in the previous 12 months. We first estimate the per-capita emigration rate, which we then feed as a prior to the ODE models. Because most households reported no departures, we modeled the household-level count of those who left $y_i$ with a zero-inflated Poisson (ZIP) model that mixes a structural-zero component with a Poisson count component:

$$y_i \sim \begin{cases} 0 & \text{with probability } \pi_i, \\ \text{Poisson}(\lambda_i) & \text{with probability } 1-\pi_i, \end{cases}$$

$$\begin{aligned}
\log(\lambda_i) &= \log(\text{PY}_i) + \beta_0 + \beta_s s_i + \alpha_{n(i)} + \eta_i, \\
\text{logit}(\pi_i) &= \gamma_0 + \gamma_s s_i + \delta_{n(i)},
\end{aligned}$$

Table S3: Model parameter descriptions. Model abbreviations are 1: SIAR model, 2: two-path immunity, 3: two-path immunity and boosting

| Parameter | Model | Units | Description | Value | Source |
|---|---|---|---|---|---|
| $R_0$ | 1,2,3 | - | Basic reproduction number | inferred | |
| $\beta_A$ | 1,2,3 | - | Relative reduction in transmission potential of asymptomatic vs. symptomatic infectors | inferred | |
| $\gamma_I$ | 1,2,3 | $wk^{-1}$ | Rate of loss of infectious state | 1/(2 days) | |
| $\mu_I$ | 1,2,3 | $wk^{-1}$ | Death rate from symptomatic infection | 0 | ignored |
| $\omega$ | 1 | $wk^{-1}$ | Rate of loss of immune protection | inferred | |
| $\omega_1$ | 2,3 | $wk^{-1}$ | Rate of loss of immune protection following asymptomatic infection | inferred | |
| $\omega_2$ | 2,3 | $wk^{-1}$ | Rate of loss of immune protection following symptomatic infection | inferred | |
| $\phi$ | 1 | - | Immune reduction of infection probability (leaky immunity) | inferred | |
| $\phi_1$ | 2,3 | - | Immune reduction of infection probability following asymptomatic infection | inferred | |
| $\phi_2$ | 2,3 | - | Immune reduction of infection probability following symptomatic infection | inferred | |
| $\mu_B$ | 1,2,3 | $wk^{-1}$ | Population birth rate | 1/(57 years) | survey |
| $\mu_{\mathcal{I}}$ | 1,2,3 | $wk^{-1}$ | Immigration rate | inferred | |
| $\mu_{\mathcal{E}}$ | 1,2,3 | $wk^{-1}$ | Emigration rate | inferred | |
| $\xi$ | 1,2,3 | - | Additional influx of infected symptomatic individuals | $1e^{-4}$ | - |
| $\alpha$ | 1,2,3 | $wk^{-1}$ | Vaccination rate | inferred | |
| $\eta$ | 1,2,3 | - | Fraction of symptomatic infections seeking healthcare at study cholera treatment centers | 2% | survey |

where $\mathrm{PY}_i$ are the person-years at risk in household $i$ (current household size plus half the number of leavers, assuming departures were uniformly distributed over the year), $s_i$ is the mean-centered household size, and $\alpha_{n(i)}$, $\delta_{n(i)}$ and $\eta_i$ are neighborhood- and household-level random effects. The per-capita emigration rate was then obtained by post-stratification over the household size distribution:

$$q = \frac{\sum_i (1-\pi_i)\,\mathrm{PY}_i\, e^{\beta_0+\beta_s s_i+\alpha_{n(i)}+\eta_i}}{\sum_i \mathrm{PY}_i}.$$

Combining $q$ with a background death rate and the observed annual population growth rate yields the total inflow rate required to balance removals and growth, which we use to inform the immigration ($\mu_{\mathcal{I}}$) and emigration ($\mu_{\mathcal{E}}$) rates of the transmission model. The ZIP formulation was preferred over Poisson and hurdle alternatives by leave-one-out cross-validation.

#### S4.3 Inference

##### S4.3.1 Model likelihood

We coordinate the model with five distinct data streams: i) weekly incidence of total AWD cases, ii) the weekly joint cross-classification of tested AWD cases by RDT result and reported vaccination status, iii) total population size in Uvira, iv) vaccination coverage in three representative household surveys in 2021, 2022 and 2023, and v) 200-day seroincidence from the 2022 serological survey.

**S4.3.1.1 AWD incidence** We link the weekly incidence of AWD cases (cholera + non-cholera AWD cases), $AWD(t)$, to the weekly incidence of symptomatic cholera infections in the models, $I_7(t)$, and a health seeking behavior parameter which represents the probability $\eta$ of a symptomatic infection requiring health care and care being sought at one of the study's cholera treatment centers:

$$
\begin{aligned}
AWD(t) &\sim f(\eta I_7(t) + \tilde{I_7}, \tau),\\
I_7(t) &= \sum_{t-7}^{t} \rho\lambda(t')X(t'),\\
\tilde{I_7}(t) &= \tilde{\lambda}(t)N(t),\\
X(t) &= \begin{cases} S(t) + S_V(t) + \phi(R(t) + R_{2V}(t)) & \text{if model = SIAR,}\\ S(t) + S_V(t) + \phi_1 R_1(t) + \phi_2(R_2(t) + R_{2V}(t)) & \text{if model = two-path,}\\ S(t) + S_V(t) + \phi_1 R_1(t) + \phi_2(R_2(t) + R_{2w}(t) + R_{2V}(t) + R_{2wV}(t)) & \text{if model = two-path-boost} \end{cases},
\end{aligned}
$$

where $\lambda(t)$ is the force of infection, $X(t)$ is the total effective susceptible population which depends on the model formulation, $\tilde{I_7}$ and $\tilde{\lambda}(t)$ are respectively the weekly incidence and incidence rates of non-cholera AWD, $N(t)$ is the population at time $t$, $f$ is the observation model and $\tau$ are the parameters of the observation model. We model the incidence rate of non-cholera AWD using cubic basis splines as detailed above for the basic reproduction number. For the observation model, we test two formulations: i) a quasi-Poisson and ii) a negative-binomial distribution, with parameter $\tau$ representing over-dispersion in both.

**S4.3.1.2 Joint RDT and vaccination status** Each week a fraction of suspected AWD cases were tested using a rapid diagnostic test (RDT), and among tested cases vaccination status was ascertained by self-report. We jointly model the weekly cross-classification of tested cases by RDT result ($R^+/R^-$) and reported vaccination status ($V^+/V^-$). For weeks in which both are recorded, the four-category counts $\mathbf{n}_{RV}(t) = (n_{R^+V^+}, n_{R^+V^-}, n_{R^-V^+}, n_{R^-V^-})$ among $N_{RV}(t)$ fully-classified cases follow a multinomial distribution:

$$\mathbf{n}_{RV}(t) \sim \text{Multinomial}\big(N_{RV}(t),\, \mathbf{p}(t)\big).$$

The cell probabilities combine the prevalence of true cholera among tested cases, $x(t)$, the RDT sensitivity $\theta^+$ and specificity $\theta^-$, and the sensitivity $\theta_V^+$ and specificity $\theta_V^-$ of self-reported vaccination. Writing $\kappa^+(t) = I_7^V(t)/I_7(t)$ for the vaccinated fraction among true cholera cases and $\kappa^-(t) = N_V(t)/N(t)$ for the vaccination coverage among non-cases, the probability of reporting $V^+$ conditional on true disease status $d \in \{+, -\}$ is

$$g_d(t) = \theta_V^+ \kappa^d(t) + (1 - \theta_V^-)\big(1 - \kappa^d(t)\big),$$

and the joint-cell probabilities are

$$
\begin{aligned}
p_{R^+V^+}(t) &= \theta^+ g_+(t)\, x(t) + (1-\theta^-) g_-(t)\,(1 - x(t)),\\
p_{R^+V^-}(t) &= \theta^+ \big(1 - g_+(t)\big) x(t) + (1-\theta^-)\big(1 - g_-(t)\big)(1 - x(t)),\\
p_{R^-V^+}(t) &= (1-\theta^+) g_+(t)\, x(t) + \theta^- g_-(t)\,(1 - x(t)),\\
p_{R^-V^-}(t) &= (1-\theta^+)\big(1 - g_+(t)\big) x(t) + \theta^- \big(1 - g_-(t)\big)(1 - x(t)),
\end{aligned}
$$

where $x(t) = \eta I_7(t)/(\eta I_7(t) + \tilde{I_7}(t))$ is the prevalence of true cholera among tested cases (as for AWD incidence), $I_7^V(t)$ is the weekly incidence of symptomatic infections among vaccinated individuals, and $N_V(t)$ is the modeled vaccinated population (see Section on vaccination coverage). The RDT sensitivity and specificity are fixed at the mean of meta-analysis estimates [9]. Weeks in which only the RDT result or only vaccination status was recorded contribute the corresponding marginal binomial likelihood, obtained by summing $\mathbf{p}(t)$ over the unobserved axis. The self-reported vaccination sensitivity and specificity are propagated by marginalizing over their estimated posterior distribution and are allowed to vary over time relative to the vaccination campaign.

**S4.3.1.3 Total population size** To inform immigration and emigration rate parameters, we link modeled population size to yearly census data available for Uvira, $P(t)$, as:

$$P(t) \sim \text{log-normal}(N(t), \sigma_P),$$

where $\sigma_P$ is the standard deviation of the error on census counts, which we here assume to be 10%.

**S4.3.1.4 Vaccination coverage** We have vaccination coverage data from our representative household surveys done in 2021 (N=2,292), 2022 (N=3,579) and 2023 (N=2,864). We link the modeled vaccinated population, $N_V(t)$, to the number of individuals responding to have received at least one OCV dose, $n_{OCV_1}(t)$, among the total number of surveyed individuals $n_{\mathcal{S}}(t)$ as:

$$\begin{aligned} n_{OCV_1}(t) &\sim \text{Binomial}(w(t), n_{\mathcal{S}}(t)), \\ w(t) &= \frac{N_V(t)}{N(t)}, \\ N_V(t) &= \begin{cases} S_V(t) + I_V(t) + A_V(t) + R_{2V}(t) & \text{if model} = \text{SIAR, two-path,} \\ S_V(t) + I_V(t) + A_V(t) + R_{2V}(t) + R_{2wV}(t) & \text{if model} = \text{two-path-boost.} \end{cases} . \end{aligned}$$

**S4.3.1.5 Seroincidence** We estimated 200-day sero-incidence from the representative survey in 2022 to be 37.9%, meaning 901 infected in the past 200 days among 2,376 participants. We therefore linked modeled 200-day cumulated infections, $I_{200}^{tot}$, to the observed counts as:

$$\begin{aligned} 901 &\sim \text{Binomial}(u(t), 2376), \\ u(t) &= \frac{I_{200}^{tot}(t_{sero})}{N(t_{sero})}, \\ I_{200}^{tot}(t) &= \begin{cases} \sum_{t-200}^{t} \lambda(t')X(t'), & \text{if model} = \text{SIAR, two-path,} \\ \sum_{t-200}^{t} \lambda(t')\left[X(t') + (1-\phi_2)(R_2(t') + R_{2V}(t'))\right], & \text{if model} = \text{two-path-boost,} \end{cases} . \end{aligned}$$

where we take $t_{sero}$ to be the midpoint of the serosurvey (March 7th 2022), and $X(t)$ is defined as for the likelihood for AWD incidence. For the SIAR and two-path models seroincidence is driven by symptomatic and asymptomatic infections. For the two-path-boost model we in addition account for boosting events.

##### S4.3.2 Priors

We set the following priors:

$$
\begin{aligned}
&\text{Disease transmission model :} \\
&R_0 \sim \mathcal{N}_{[0,20]}(5, 2.5), \\
&\text{logit}(\beta_A) \sim \mathcal{N}(\text{logit}(0.2), 0.75), \\
&\log(\omega), \log(\omega_2) \sim \mathcal{N}_{[\log(1/(51\times 20)),\log(1/51)]}(\log(1/(7\times 51)), 0.35) \\
&\log(\omega_1) \sim \mathcal{N}_{[\log(1/(51\times 10)),\log(1/2)]}(\log(1/(0.75\times 51)), 0.5), \\
&\text{logit}(\phi), \text{logit}(\phi_2) \sim \mathcal{N}(\text{logit}(0.05), 0.75), \\
&\text{logit}(\phi_1) \sim \mathcal{N}(\text{logit}(0.25), 0.75), \\
&\mu_{\mathcal{I}}, \mu_{\mathcal{E}} \sim \mathcal{N}_{[0,0.001]}(1/(51\times 5), 0.0005), \\
&\log(\alpha) \sim \mathcal{N}_{[-\log(1-0.66),-\log(1-0.8)]}(-\log(1-0.67), 0.75), \\
&a_i \sim \mathcal{N}(0,1), \\
&\tilde{a_i} \sim \mathcal{N}(0,1), \\
&\text{Observation model :} \\
&1/\tau \sim \mathcal{N}_{[0,\infty)}(0,1), \\
&\text{Initial conditions :} \\
&I(0) \sim \mathcal{N}_{[0,1]}(1e^{-5}, 1e^{-3}), \\
&A(0) \sim \mathcal{N}_{[0,1]}(1e^{-4}, 1e^{-3}), \\
&\text{SIAR model :} \\
&R(0) \sim \mathcal{N}_{[0,1]}(0.85, 0.1), \\
&1 = S(0) + I(0) + A(0) + R(0), \\
&\text{two-path model :} \\
&R_1(0) \sim \mathcal{N}_{[0,1]}(0.1, 0.2), \\
&R_2(0) \sim \mathcal{N}_{[0,1]}(0.75, 0.25), \\
&1 = S(0) + I(0) + A(0) + R_1(0) + R_2(0), \\
&\text{two-path-boost model :} \\
&R_1(0) \sim \mathcal{N}_{[0,1]}(0.1, 0.2), \\
&R_2(0) \sim \mathcal{N}_{[0,1]}(0.45, 0.25), \\
&R_{2w}(0) \sim \mathcal{N}(0.45, 0.25), \\
&1 = S(0) + I(0) + A(0) + R_1(0) + R_2(0) + R_{2w}(0),
\end{aligned}
$$

where $\mathcal{N}_{[a,b]}$ denotes a truncated normal distribution with lower bound $a$ and upper bound $b$, $\tilde{a_i}$ are the B-spline coefficients of the non-cholera AWD incidence rate, and the initial conditions for each model are defined as a simplex on the scaled population (i.e. $N(0) = 1$);

#### S4.4 Counterfactual analysis

##### S4.4.1 Analysis setup

To perform counterfactual analysis we modified the vaccination and population turnover rate parameters running the model on draws of the posterior distribution of the time varying $R_0$ as inferred from the data.

##### S4.4.2 Model ensembling

To build the model ensemble we sampled from the stacked posterior draws of all model formulations. Each model posterior draw was assigned a weight using the stacking weights, and draws were sampled without replacement based on these weights [10].

#### S4.5 Sensitivity analyses

We conducted two pre-specified sensitivity analyses to assess the robustness of the main findings to alternative assumptions about key data inputs. Both sensitivity analyses used the same four model formulations as the main analysis (two-path, two-path with R1V compartment, two-path with immune-boosting, and two-path with immune-boosting and R1V compartment, all with negative-binomial observation model), with the same model ensemble construction procedure (stacking weights excluding the simplified single-recovery model).

##### S4.5.1 Health seaking at cholera treamtment center sensitivity

The main analysis assumed a constant health-seeking fraction of 2% throughout the study period (i.e., 2% of symptomatic cholera cases were estimated to present to a health facility and be captured in surveillance). However, active case finding and enhanced data collection under this surveillance study (which began in January 2020) may have increased case ascertainment. To assess sensitivity to this assumption, we re-ran the full analysis with the health-seeking fraction increased by a factor of 1.25 from January 2020 onwards (effective health-seeking fraction of 2.5% during the enhanced surveillance period), while retaining the same 2% fraction for the pre-study period (2017–2019). All other model parameters were kept identical to the main analysis.

##### S4.5.2 Seroincidence sensitivity

The main analysis used a 200-day seroincidence of 37.9%, derived from the April 2022 seroprevalence survey, as an informative likelihood data point constraining the inferred level of population immunity. To assess sensitivity to this estimate, we re-ran the analysis targeting a seroincidence of 60% (high) or 15% (low), reflecting uncertainty in the survey-derived estimate and the possibility of higher or lower cumulative population exposure than captured by the survey. All other model parameters were kept identical to the main analysis.

##### S4.5.3 Results

Stacked posterior estimates of vaccine impact for both sensitivity analyses are presented alongside the main analysis results in Supplementary Table S7.

### S5 Supplementary Figures

#### Figure S2: Seasonality

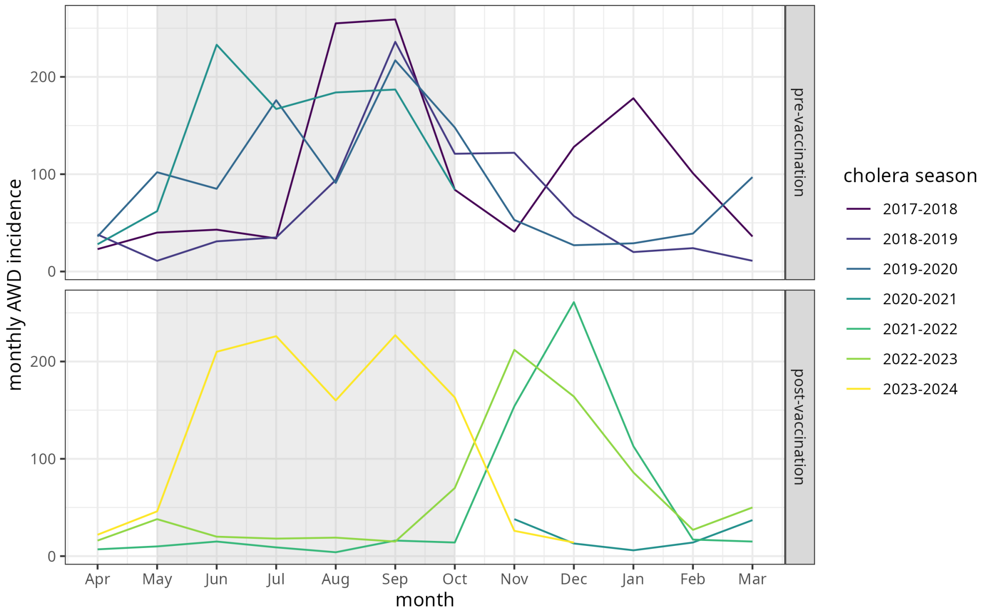

Figure S2: Seasonality of cholera in Uvira, DRC. Cholera years are defined as starting in April each year. Gray windows (May-October) indicate typical periods of seasonal cholera peaks.

#### Figure S3: Wavelet analysis

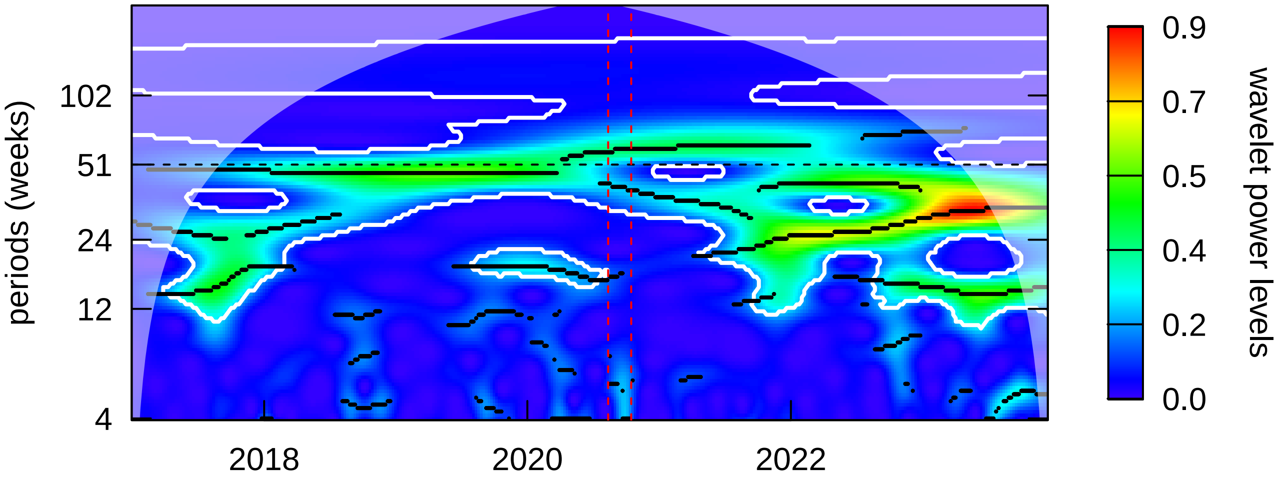

Figure S3: Wavelet analysis. Continuous wavelet power spectrum of the weekly reconstructed cholera incidence series using the Morlet wavelet. Warmer colours indicate higher wavelet power. The horizontal dashed line marks the annual (51-week) period and the vertical dashed red lines the two rounds of the 2020 OCV campaign.

#### Figure S4: Additional model fidelity plots

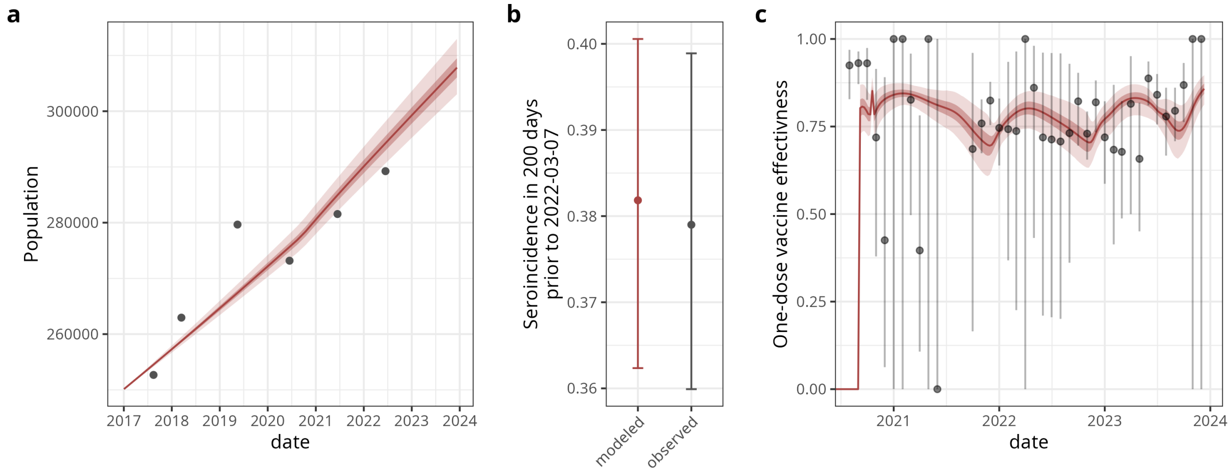

Figure S4: Additional model fidelity plots. Plots are given for the best-fitting model (immune boosting process model and negative-binomial observation model) for a) population of Uvira, DRC, b) 200-day seroincidence on March 7th 2022, and c) vaccine effectiveness. Red lines and dots indicate the mean of 2,000 HMC model posterior draws, and red ribbons and errorbars the 95% CrIs. Gray dots and errorbars indicate in a) population census counts in Uvira, in b) the mean and 95% CIs of 200-day seroincidence as detailed in the Methods section of the main, and in c) the mean and 95% CIs of monthly vaccine effectiveness estimated using the screening method on available surveillance data as detailed in Supplementary Section S3 (dates with missing estimates correspond to periods with limited sample size).

#### Figure S5: Model parameters

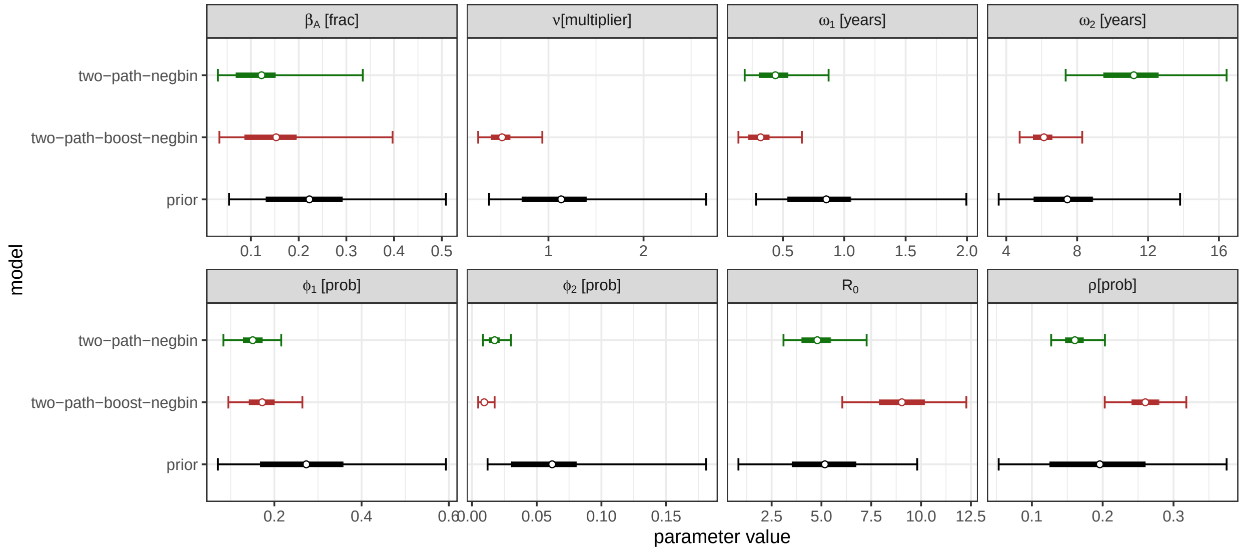

Figure S5: Parameter estimates for the two best fitting models (two-path and two-path-boost models with negative-binomial observation model). Dots indicate the mean of 2,000 HMC posterior draws for the two-path (red) and two-path-boost (black) models, thick bars the 50% CrIs, and whiskers the 95% CrIs. Parameters are described in Supplementary Table S3.

#### Figure S6: Implied protection after infection/vaccination

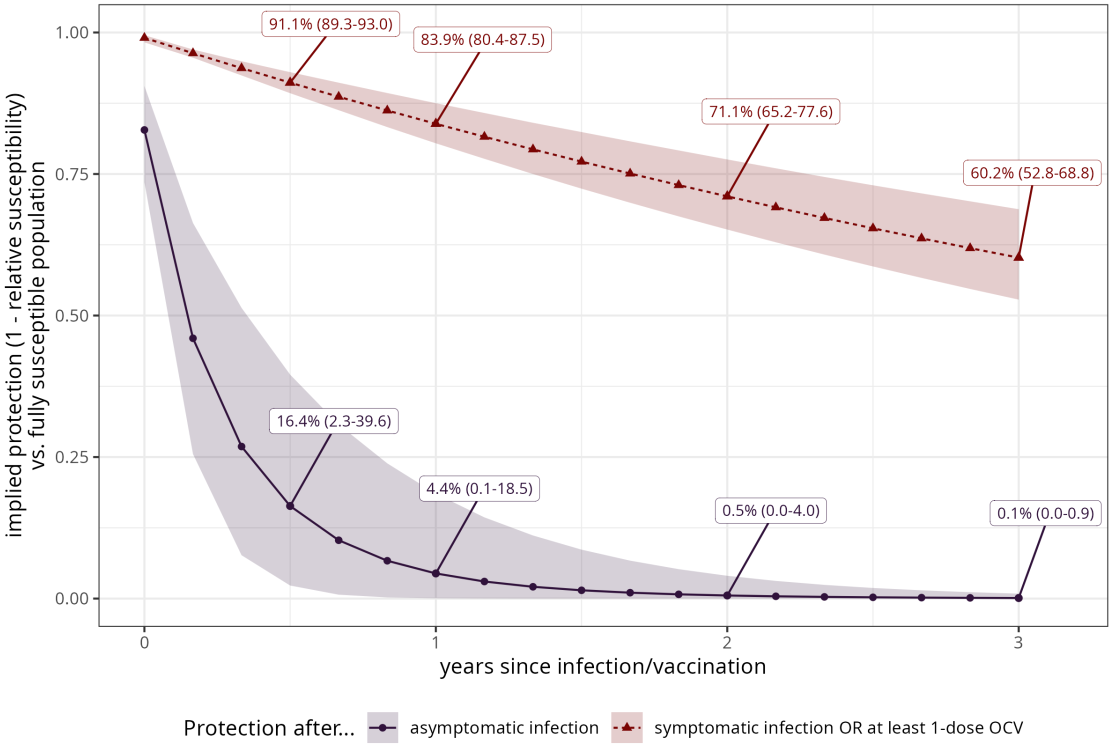

Figure S6: Estimated immune protection in comparison to a fully suscepltible population after asymptomatic infection vs. symptomatic or at least 1-dose OCV (assumed equal in the models). Lines indicate the mean of 2,000 HMC posterior draws and ribbons the 95% CrIs.

#### Figure S7: Basic reproduction number estimates

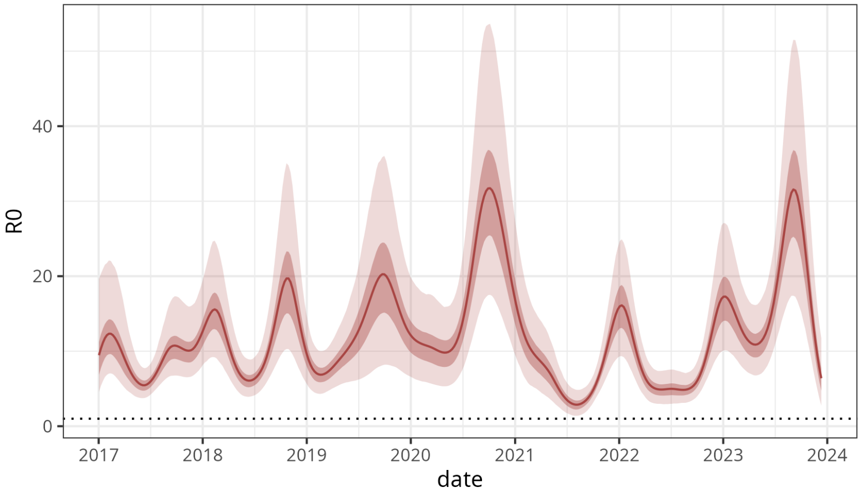

Figure S7: Estimate of the time-varying basic reproduction number for the best fitting model (immune boosting process model and negative-binomial observation model). The line indicates the mean of 2,000 HMC posterior draws and ribbons the 95% CrIs.

#### Figure S8: All basic reproduction number estimates

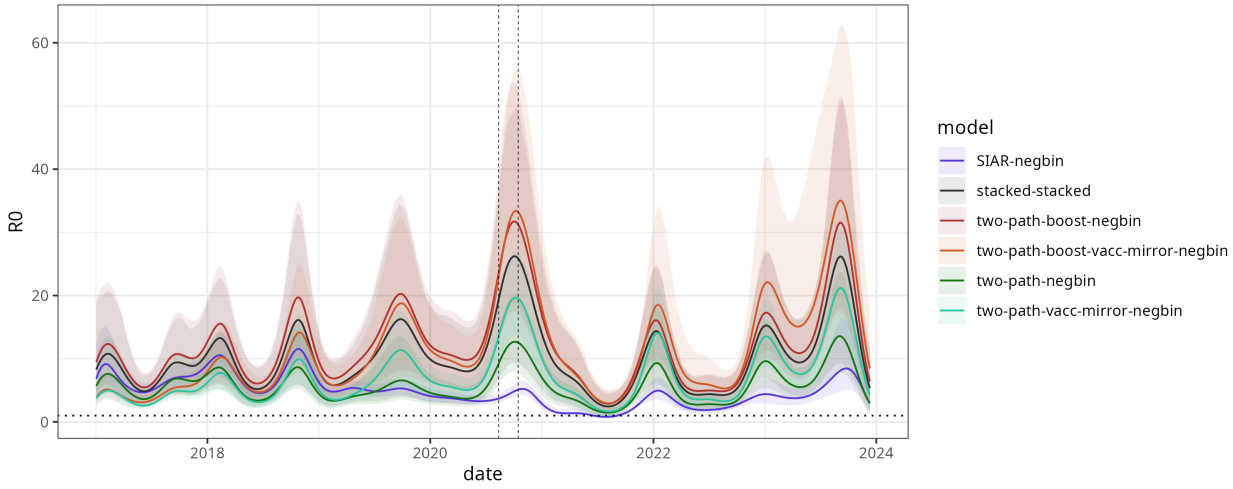

Figure S8: Estimate of the time-varying basic reproduction number for all models. The line indicates the mean of 2,000 HMC posterior draws and ribbons the 95% CrIs.

#### Figure S9: All states

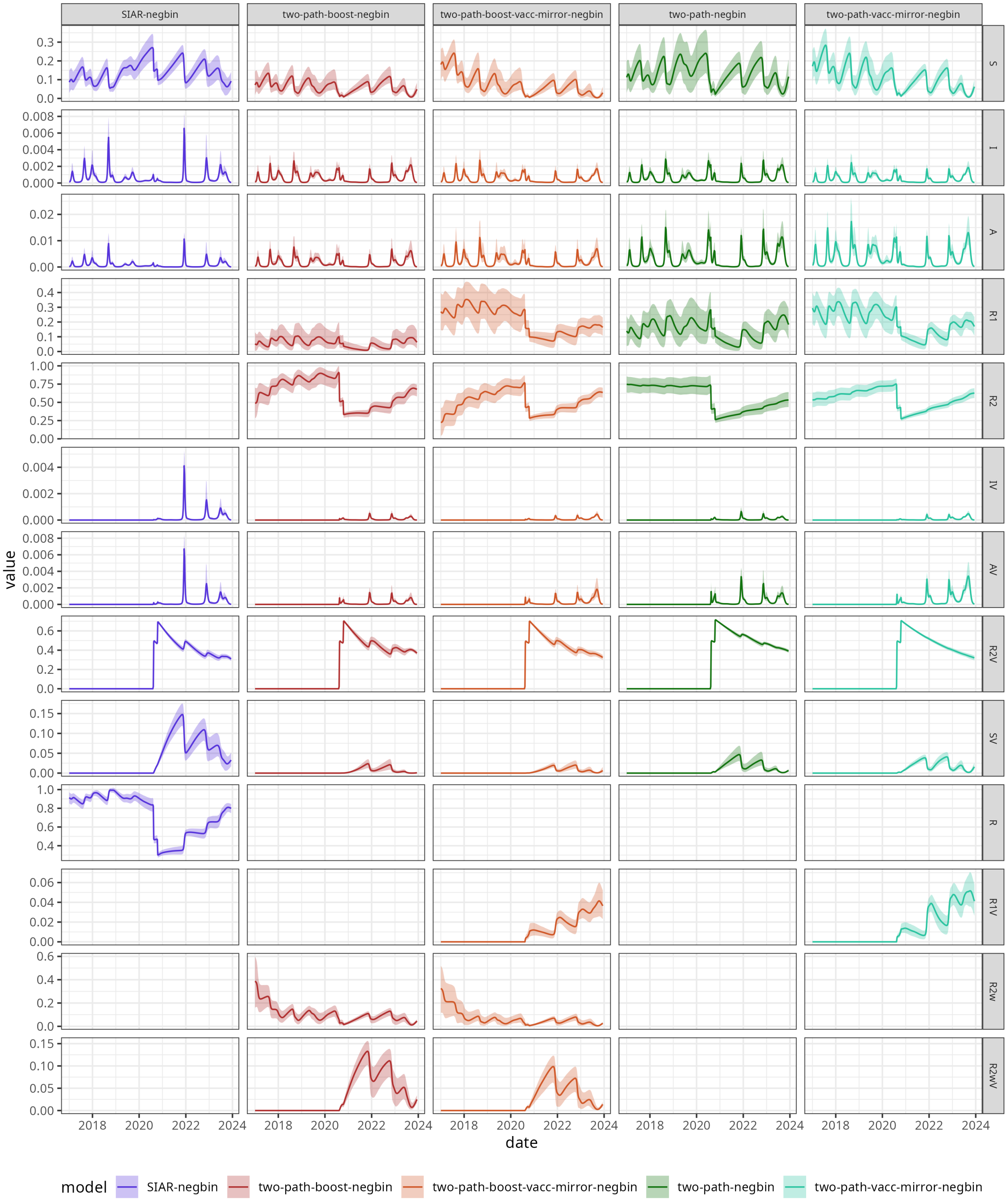

Figure S9: Estimated trajectories of all model compartments (states) for all models. Lines indicate the mean of 2,000 HMC posterior draws and ribbons the 95% CrIs.

#### Figure S10: Averted symptomatic cases per OCV dose

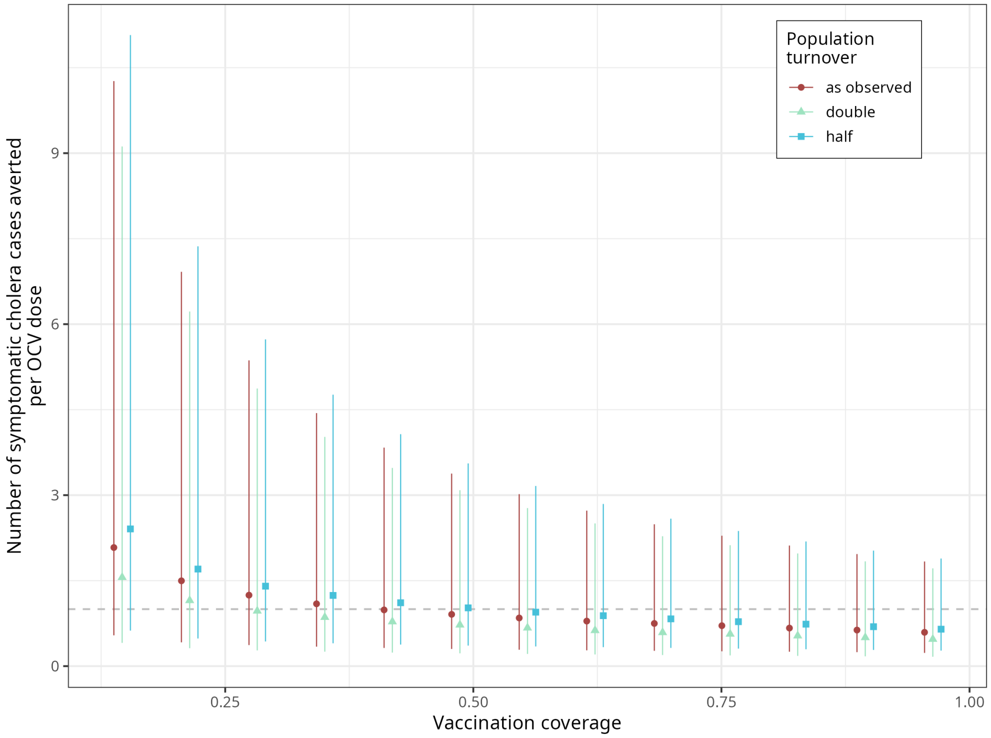

Figure S10: Estimated number of averted symptomatic cases per OCV dose in the three years post vaccination. Dots indicate the median of 2,000 HMC posterior draws and ribbons the 95% CrIs.

### S6 Supplementary Tables

#### Table S4: Model comparison results.

Table S4: Model comparison. SIAR model was dropped from model ensemble therefore does not have weight.

| Model | ELPD diff | SE | Rank | Significant | Weight |
|---|---|---|---|---|---|
| **two-path-boost-negbin** | 0.0 | 0.0 | 1 | FALSE | 0.686 |
| **two-path-negbin** | -8.8 | 5.9 | 2 | FALSE | 0.245 |
| **two-path-vacc-mirror-negbin** | -47.5 | 12.4 | 3 | TRUE | 0.069 |
| **two-path-boost-vacc-mirror-negbin** | -56.5 | 12.0 | 4 | TRUE | 0.000 |
| **SIAR-negbin** | -212.3 | 38.1 | 5 | TRUE | NA |

#### Table S5: Parameter posterior summaries and convergence diagnostics.

Table S5: Posterior summaries and MCMC convergence diagnostics for free parameters of each model. Mean and 95% credible intervals (CrI) are on the natural scale (back-transformed from the sampling parameterisation). $\hat{R}$ is the potential-scale-reduction factor; values $\leq 1.01$ indicate good mixing. $\mathrm{ESS}_{\mathrm{bulk}}$ is the bulk effective sample size.

| Parameter | Unit | Mean [95% CrI] | $\hat{R}$ | $\mathrm{ESS}_{\mathrm{bulk}}$ |
|---|---|---|---|---|
| **two-path-boost-negbin** | | | | |
| $R_0$ | - | 9.04 [6.05, 12.3] | 1.001 | 1353 |
| $\beta_A$ | frac | 0.153 [0.0337, 0.397] | 1.004 | 1752 |
| $\lambda$ | rate | 1.08 [1.08, 1.09] | 1.001 | 2336 |
| $\mu_B$ | years | 5.1 [4.78, 5.45] | 1.001 | 2237 |
| $\mu_D$ | years | 5.24 [4.89, 5.6] | 1.001 | 2218 |
| $\nu$ | multiplier | 0.512 [0.261, 0.936] | 1.003 | 1425 |
| $\omega_1$ | years | 0.318 [0.138, 0.655] | 1.001 | 1237 |
| $\omega_2$ | years | 6.12 [4.76, 8.29] | 1.006 | 1008 |
| $\phi_1$ | prob | 0.172 [0.0945, 0.264] | 1.002 | 1348 |
| $\phi_2$ | prob | 0.00946 [0.00478, 0.0175] | 1.007 | 854 |
| $\rho_I$ | prob | 0.26 [0.203, 0.318] | 1.001 | 1831 |
| **two-path-boost-vacc-mirror-negbin** | | | | |
| $R_0$ | - | 8.09 [5.41, 11.4] | 1.005 | 736 |
| $\beta_A$ | frac | 0.103 [0.0287, 0.244] | 1.001 | 1451 |
| $\lambda$ | rate | 1.08 [1.08, 1.1] | 1.005 | 2436 |
| $\mu_B$ | years | 4.77 [4.5, 5.09] | 1.000 | 1790 |
| $\mu_D$ | years | 4.92 [4.61, 5.26] | 1.001 | 1869 |
| $\nu$ | multiplier | 0.774 [0.29, 1.81] | 1.002 | 2188 |
| $\omega_1$ | years | 1.18 [0.455, 2.48] | 1.007 | 512 |
| $\omega_2$ | years | 7.98 [5.76, 12.5] | 1.013 | 651 |
| $\phi_1$ | prob | 0.0978 [0.0517, 0.16] | 1.004 | 657 |
| $\phi_2$ | prob | 0.00579 [0.00194, 0.0116] | 1.009 | 537 |
| $\rho_I$ | prob | 0.199 [0.146, 0.268] | 1.003 | 776 |
| **two-path-negbin** | | | | |
| $R_0$ | - | 4.79 [3.1, 7.27] | 1.008 | 590 |
| $\beta_A$ | frac | 0.122 [0.0308, 0.334] | 1.003 | 1569 |
| $\lambda$ | rate | 1.08 [1.08, 1.1] | 1.001 | 2053 |
| $\mu_B$ | years | 5.15 [4.83, 5.51] | 1.001 | 2060 |
| $\mu_D$ | years | 5.28 [4.95, 5.66] | 1.001 | 1954 |
| $\omega_1$ | years | 0.439 [0.189, 0.872] | 1.005 | 1036 |
| $\omega_2$ | years | 11.2 [7.35, 16.4] | 1.001 | 1335 |
| $\phi_1$ | prob | 0.15 [0.0831, 0.216] | 1.002 | 1270 |
| $\phi_2$ | prob | 0.0175 [0.00843, 0.0301] | 1.008 | 522 |
| $\rho_I$ | prob | 0.161 [0.127, 0.203] | 1.002 | 1319 |
| **two-path-vacc-mirror-negbin** | | | | |
| $R_0$ | - | 5.5 [3.85, 7.43] | 1.000 | 1887 |
| $\beta_A$ | frac | 0.0786 [0.0247, 0.177] | 1.002 | 2869 |
| $\lambda$ | rate | 1.08 [1.08, 1.1] | 0.999 | 2355 |
| $\mu_B$ | years | 4.71 [4.42, 5.02] | 1.002 | 3174 |
| $\mu_D$ | years | 4.84 [4.53, 5.17] | 1.001 | 3170 |
| $\omega_1$ | years | 0.568 [0.295, 0.954] | 1.000 | 1555 |
| $\omega_2$ | years | 17.7 [14.3, 19.9] | 1.000 | 2010 |
| $\phi_1$ | prob | 0.141 [0.0924, 0.202] | 1.000 | 1535 |
| $\phi_2$ | prob | 0.0035 [0.00136, 0.00705] | 1.000 | 1787 |
| $\rho_I$ | prob | 0.123 [0.101, 0.148] | 1.000 | 2224 |

#### Table S6: Averted cases by year post-vaccination.

Table S6: Stacked posterior estimates of the proportion and number of averted cholera cases by year following the August 2020 OCV campaign and over the full post-vaccination period. Values are median [95% CrI] from the stacked model ensemble (counter-factual scenario 1: no vaccination). The third year covers 16 months (August 2022–December 2023).

| Period | % cases averted | Averted medically-attended cases | Averted community symptomatic cases |
|---|---|---|---|
| Year 1 (Aug 2020–Aug 2021) | 79.5% (66.9-94.3) | 834 (352-4,362) | 41,728 (17,554-218,150) |
| Year 2 (Aug 2021–Aug 2022) | 45.1% (27.9-67.1) | 250 (118-738) | 12,488 (5,876-36,914) |
| Year 3 (Aug 2022–Dec 2023) | 47.8% (11.2-76.9) | 1,234 (140-4,852) | 61,720 (6,978-242,562) |
| Overall (Aug 2020–Dec 2023) | 56.3% (34.3-81.2) | 2,472 (878-8,356) | 123,586 (43,916-417,844) |

##### S6.1 Table S7: Stacked impact estimates: sensitivity analyses.

Table S7: Stacked posterior estimates of averted cholera cases over the full post-vaccination period (August 2020–December 2023), main analysis and sensitivity analyses. Values are median [95% CrI] from the stacked model ensemble (counter-factual scenario 1: no vaccination).

| Analysis | % cases averted | Averted medically-attended cases | Averted community symptomatic cases |
|---|---|---|---|
| **Main analysis** | | | |
| Main analysis | 56.3% (34.4-81.3) | 2,472 (878-8,356) | 123,586 (43,916-417,844) |
| **Sensitivity analysis** | | | |
| Higher health seeking (x1.25) | 47.0% (31.4-72.6) | 1,464 (756-5,806) | 58,584 (30,260-232,230) |
| Higher seroincidence (60%) | 65.8% (32.5-87.9) | 3,896 (814-15,586) | 194,830 (40,702-779,322) |
| Lower seroincidence (15%) | 37.3% (26.0-52.9) | 1,062 (586-2,096) | 53,116 (29,306-104,848) |

### References

[1] CP Farrington. "Estimation of vaccine effectiveness using the screening method". In: *International journal of epidemiology* 22.4 (1993), pp. 742–746.

[2] Aaron A King et al. "Inapparent infections and cholera dynamics". In: *Nature* 454.7206 (2008), pp. 877–880.

[3] Elizabeth C Lee et al. "Achieving coordinated national immunity and cholera elimination in Haiti through vaccination: a modelling study". In: *The Lancet Global Health* 8.8 (2020), e1081–e1089.

[4] Andrew S Azman et al. "The impact of a one-dose versus two-dose oral cholera vaccine regimen in outbreak settings: a modeling study". In: *PLoS Medicine* 12.8 (2015), e1001867.

[5] Adam Le et al. "The Impact of Infection-Derived Immunity on Disease Dynamics". In: *Journal of Mathematical Biology* 83.6 (Nov. 2021), p. 61. ISSN: 1432-1416. DOI: 10.1007/s00285-021-01681-4. (Visited on 02/19/2024).

[6] Odo Diekmann, Johan Andre Peter Heesterbeek, and Johan AJ Metz. "On the definition and the computation of the basic reproduction ratio $R_0$ in models for infectious diseases in heterogeneous populations". In: *Journal of Mathematical Biology* 28.4 (1990), pp. 365–382.

[7] Judith A Bouman et al. "Bayesian workflow for time-varying transmission in stratified compartmental infectious disease transmission models". In: *PLoS computational biology* 20.4 (2024), e1011575.

[8] Aybüke Koyuncu et al. "Challenges with achieving and maintaining oral cholera vaccine coverage: insights from serial cross-sectional representative surveys in a cholera-endemic community in the Democratic Republic of the Congo". In: *BMJ Public Health* 3.1 (2025).

[9] Basilua Andre Muzembo et al. "Accuracy of cholera rapid diagnostic tests: a systematic review and meta-analysis". In: *Clinical Microbiology and Infection* 28.2 (2022), pp. 155–162.

[10] Aki Vehtari and Jonah Gabry. "Bayesian Stacking and Pseudo-BMA weights using the loo package". In: *Version loo* 2.0 (2019).
